# A Drift-Oriented Adaptive Framework for Concept Drift Detection in Large-Scale Internet-of-Medical-Things Data Streams

**DOI:** 10.1101/2024.02.16.24302969

**Authors:** Vikash Maheshwari, Nurul Aida Bt. Osman, Hanita Daud, Angelina Prima Kurniati, Wan Nur Syahidah Bt. Wan Yusoff

**Affiliations:** Computer and Information Science Department, Universiti Teknologi PETRONAS, Bandar Seri Iskandar 32610, Perak, Malaysia; Centre for Research in Data Science (CERDAS), Universiti Teknologi PETRONAS, Seri Iskandar 32610, Malaysia; Department of Fundamental and Applied Sciences, Universiti Teknologi PETRONAS, Seri Iskandar 32610, Malaysia; School of Computing, Telkom University, Indonesia; Centre for Mathematical Sciences, Universiti Malaysia Pahang, Malaysia

**Author notes:** Correspondence (V.M.). (V.M). These authors contributed equally to this work. **Conceptualization:** Vikash Maheshwari. **Data curation:** Vikash Maheshwari. **Formal analysis:** Vikash Maheshwari, ¶Nurul Aida Bt. Osman, ¶Hanita Daud. **Funding acquisition:** Nurul Aida Bt. Osman, Hanita Daud, Angelina Prima Kurniati, and Wan Nur Syahidah Bt. Wan Yusoff. **Methodology:** Vikash Maheshwari, ¶Nurul Aida Bt. Osman. **Writing – original draft:** Vikash Maheshwari. **Writing – review & editing:** Vikash Maheshwari, ¶Nurul Aida Osman, Angelina Prima Kurniati, and Wan Nur Syahidah Bt. Wan Yusoff. **Supervision:** Nurul Aida Osman. All authors have read and agreed to the published version of the manuscript.

**Keywords:** Internet of Medical Things (IoMT), concept drift, healthcare systems, IoT, AEF-CDA

## Abstract

**Background:** The rise of the Internet-of-Medical-Things (IoMT) and smart devices has led to a substantial increase in extensive data streams in the healthcare domain. The interconnected nature of medical devices introduces dynamic and evolving data patterns. However, this dynamism poses a significant challenge known as Concept Drift, particularly crucial in the medical field. Concept Drift reflects the inherent instability in data patterns over time. In medical applications, this challenge intensifies as sensors must seamlessly transition from general healthcare monitoring to handling critical situations like emergency ICU operations. The complexity deepens due to imbalanced data distributions inherent in e-health scenarios.

**Methods:** The study introduces an Adaptive Ensemble Framework (AEF-CDA) designed to detect and adapt to concept drift in large-scale medical data streams from IoMT. The framework incorporates adaptive data preprocessing, a novel drift-centric adaptive feature selection approach, the learning of base models, and the selection of models adapted to concept drift. Additionally, an online ensemble model is integrated to enhance concept drift adaptation.

**Results:** The proposed AEF-CDA framework is evaluated using three public IoMT and IoT datasets. The experimental results demonstrate its superiority over contemporary methods, achieving a remarkable accuracy of 99.64% with a precision of 99.39%. These metrics surpass the performance of other approaches in the simulation.

**Conclusion:** In conclusion, the research presents a robust solution in the form of the adaptive ensemble framework (AEF-CDA) to effectively address the challenges posed by concept drift in IoMT data streams. The demonstrated high accuracy and precision underscore the framework’s efficacy, highlighting its potential significance in the dynamic landscape of medical data analysis.

## 1 Introduction

The Internet of Things (IoT) has recently transformed how people live their lives by combining low-cost sensors with the Internet. This integration has made it possible to collect data on physical world events and manage physical infrastructure [1]. IoT is also applicable in areas including automation in factories, healthcare, and transportation. The use of IoT in healthcare is now thought to be one of the most popular subjects for research. According to [2], the Internet of Medical Things (IoMT) is significantly contributing to the healthcare industry by making electronic devices more accurate, reliable, and efficient. Considering the current pandemic situation, visiting a physician for minor health concerns poses a substantial risk to individuals. As a result, the adoption of IoMT devices facilitates convenient monitoring of daily health records, thereby allowing individuals to take the necessary precautions on their own.

According to a recent survey [3], the adoption of IoT in numerous areas of life has occurred at varying rates. As per a report, it is projected that around 41.6 billion devices within the Internet of Things (IoT) will be operational by the year 2025, leading to the creation of an estimated 79.4 zettabytes (ZB) of data. Furthermore, the revenue generated by IoT is expected to rise from $5.6 billion in 2016 to around $27 billion by 2024. In addition to enhancing healthcare monitoring, management, and medical procedures, the Internet of Medical Things (IoMT) can lead to improved human well-being and better quality of life [4][5]. By 2030, there will be a significant shortage of healthcare workers, according to predictions from the World Health Organization (WHO). It has resulted in efforts to develop cost-effective smart healthcare infrastructure and solutions, aimed at preventing diseases, reducing healthcare expenditures, and improving the quality of life for all [6]. Table 1 outlines the different categories and characteristics of medical devices utilized on the Internet of Medical Things (IoMT) system.

**Table 1.** The various categories and attributes of medical devices employed in the IoMT system.

| Ref. | Type of Device | Placement | Risk Level | Description |
| --- | --- | --- | --- | --- |
| [7] | Stationary | In the hospitals | Low | MRI and CT-Scan medical image processing equipment |
| [8] | Wearable | On the body of a human | Low | Smartwatches and fitness equipment |
| [9] | Implantable | Throughout human tissues | High | The cardiac pacemaker, the insulin pump, and deep brain implants |
| [10] | Ambient | Outside the body of a human | Low | Monitoring equipment for the elderly in smart homes |

**Table 2.**
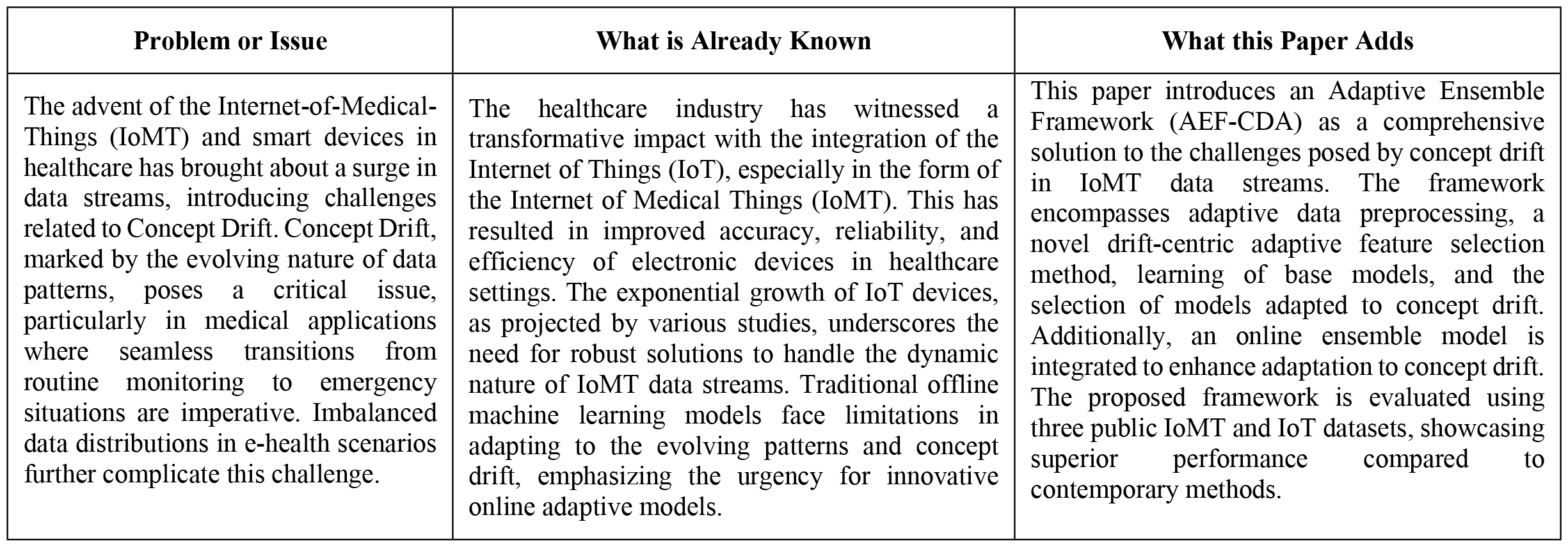
Statement of Significance.

| Problem or Issue | What is Already Known | What this Paper Adds |
| --- | --- | --- |
| The advent of the Internet-of-Medical-Things (IoMT) and smart devices in healthcare has brought about a surge in data streams, introducing challenges related to Concept Drift. Concept Drift, marked by the evolving nature of data patterns, poses a critical issue, particularly in medical applications where seamless transitions from routine monitoring to emergency situations are imperative. Imbalanced data distributions in e-health scenarios further complicate this challenge. | The healthcare industry has witnessed a transformative impact with the integration of the Internet of Things (IoT), especially in the form of the Internet of Medical Things (IoMT). This has resulted in improved accuracy, reliability, and efficiency of electronic devices in healthcare settings. The exponential growth of IoT devices, as projected by various studies, underscores the need for robust solutions to handle the dynamic nature of IoMT data streams. Traditional offline machine learning models face limitations in adapting to the evolving patterns and concept drift, emphasizing the urgency for innovative online adaptive models. | This paper introduces an Adaptive Ensemble Framework (AEF-CDA) as a comprehensive solution to the challenges posed by concept drift in IoMT data streams. The framework encompasses adaptive data preprocessing, a novel drift-centric adaptive feature selection method, learning of base models, and the selection of models adapted to concept drift. Additionally, an online ensemble model is integrated to enhance adaptation to concept drift. The proposed framework is evaluated using three public IoMT and IoT datasets, showcasing superior performance compared to contemporary methods. |

The Internet of Medical Things (IoMT) represents a subset within the broader category of the Internet of Things (IoT), functioning as a bridge connecting the Internet with the healthcare industry. The Internet of Medical Things (IoMT) represents the future of modern healthcare, which will allow healthcare professionals to monitor and interconnect medical devices via the internet. As part of the Internet of Things, the IoMT integrates a wide range of products and platforms, including embedded devices, elder-care wearables for observing [11], and internet-connected clinical equipment for remote surgery [7]. In addition to tracking heart rates and movements, smart watches are available. Likewise there are contact lenses made specifically to measure blood sugar levels.

Figure 1. depicts the integration of several medical-related technologies to form the IoMT (Internet of Medical Things). A high-speed stream of medical data is continuously generated by IoMT devices, requiring real-time analysis. IoMT data distributions evolve with time, leading to concept drift in IoMT device applications in the real world. The concept drift will pose significant challenges in the development of IoMT systems since it can lead to a gradual degradation of their performance as a result of changing data distributions [12]. In addition, it is challenging to adapt to the continuously changing environment due to the evolving behaviors of IoT attacks. Thus, in the medical domain, timely analysis is not only important, but also the detection and adaptation of concept drift is essential. Furthermore, traditional offline machine learning (ML) models lack the capability to effectively manage concept drift [12]. This underscores the need for the creation of innovative online adaptive models designed to identify and adjust to concept drift within data streams from the Internet of Medical Things (IoMT).

**Figure 1.**
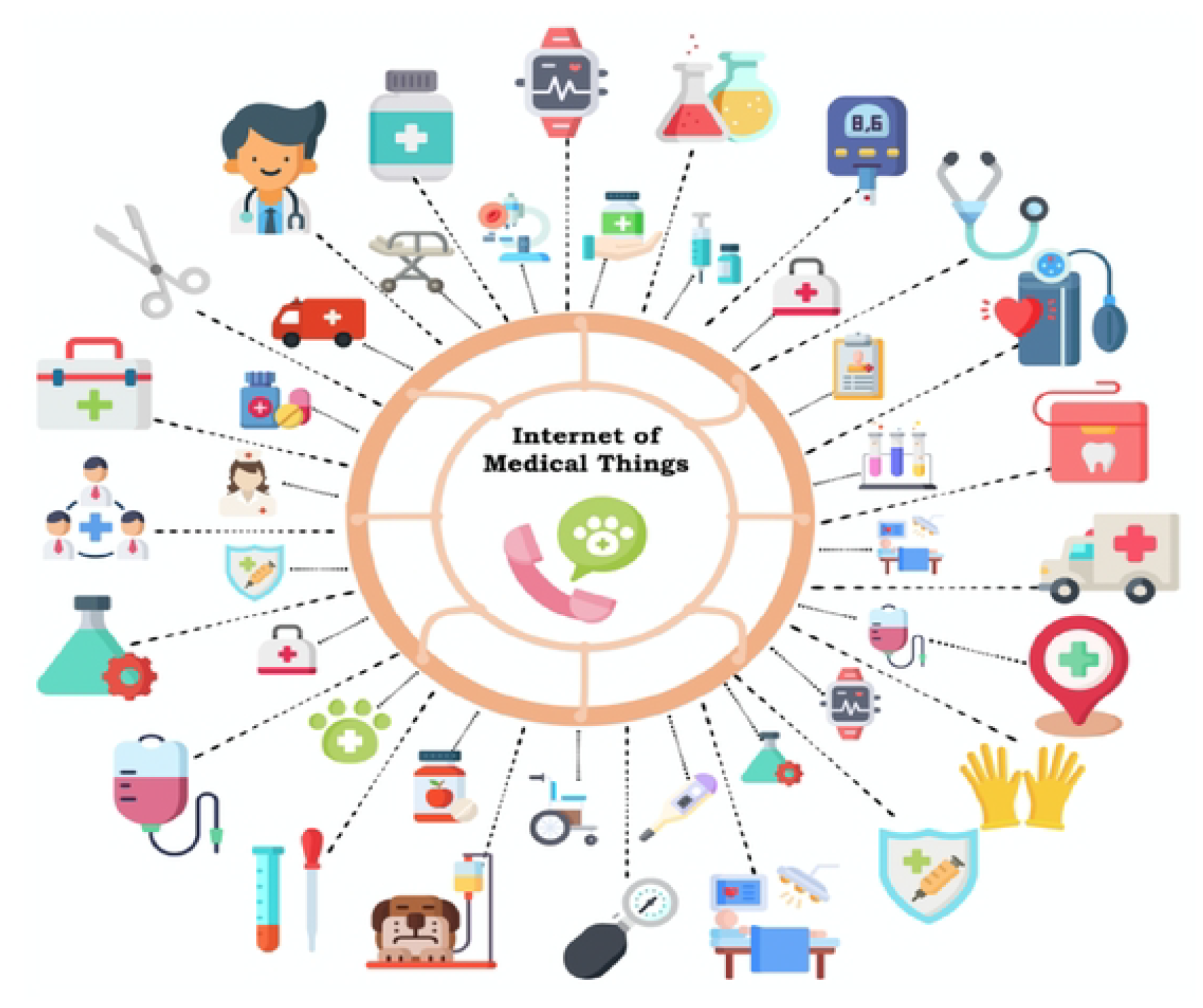
An overview of the Internet of Medical Things (IoMT) infrastructure.

This paper proposes a drift-oriented adaptive ensemble framework for the real-time identification and adaptation of concept drift in IoMT data stream. The outlined framework comprises multiple phases, encompassing procedures such as data pre-processing, normalization, and the establishment of an adaptive ensemble framework. To assess the efficacy of the proposed framework and tackle security concerns associated with the Internet of Medical Things (IoMT), various benchmark datasets are employed. These datasets encompass the WUSTL EHMS-IoMT medical dataset, IoTID20, and CICIDS2017 datasets.

Following is a summary of the contributions of the proposed study:

- It suggests an novel and thorough framework for identifying and adapting to concept drift within the data stream of the Internet of Medical Things (IoMT).
- It introduces Drift-Adaptive Dynamic Feature Selection (DA-FS), a unique method for selecting features, specifically designed for analyzing medical data streams that face challenges related to concept drift.
- To investigate different health domains within the IoMT framework and their utilization in the intelligent healthcare system, encompassing the types of sensors employed for each specific domain.
- We chose accuracy as an assessment metric to evaluate our framework’s performance and obtained the highest result of 99.64 %.

The paper is organized into the following sections: In Section 2, the architecture of IoMT is detailed, while Section 3 offers an overview of pertinent literature regarding concept drift detection and adaptation. Section 4 introduces the proposed methodology for tackling the concept drift issue. Performance evaluation is discussed in Section 5. The paper concludes with a summary in Section 6.

## 2 Statement of Significance

## 3 Internet of Medical Things (IoMT) Architecture

Figure 2 depicts the three levels of the IoMT architecture: the items layer, the fog layer, and the cloud layer [13]. This architectural design is a variant of the original framework provided in [14]. With this configuration, healthcare professionals may interact directly with one another via the router that connects the Thing layer and the Fog layer, as well as via the local processing servers placed at the Fog layer. Each layer is described below:

**Figure 2.**
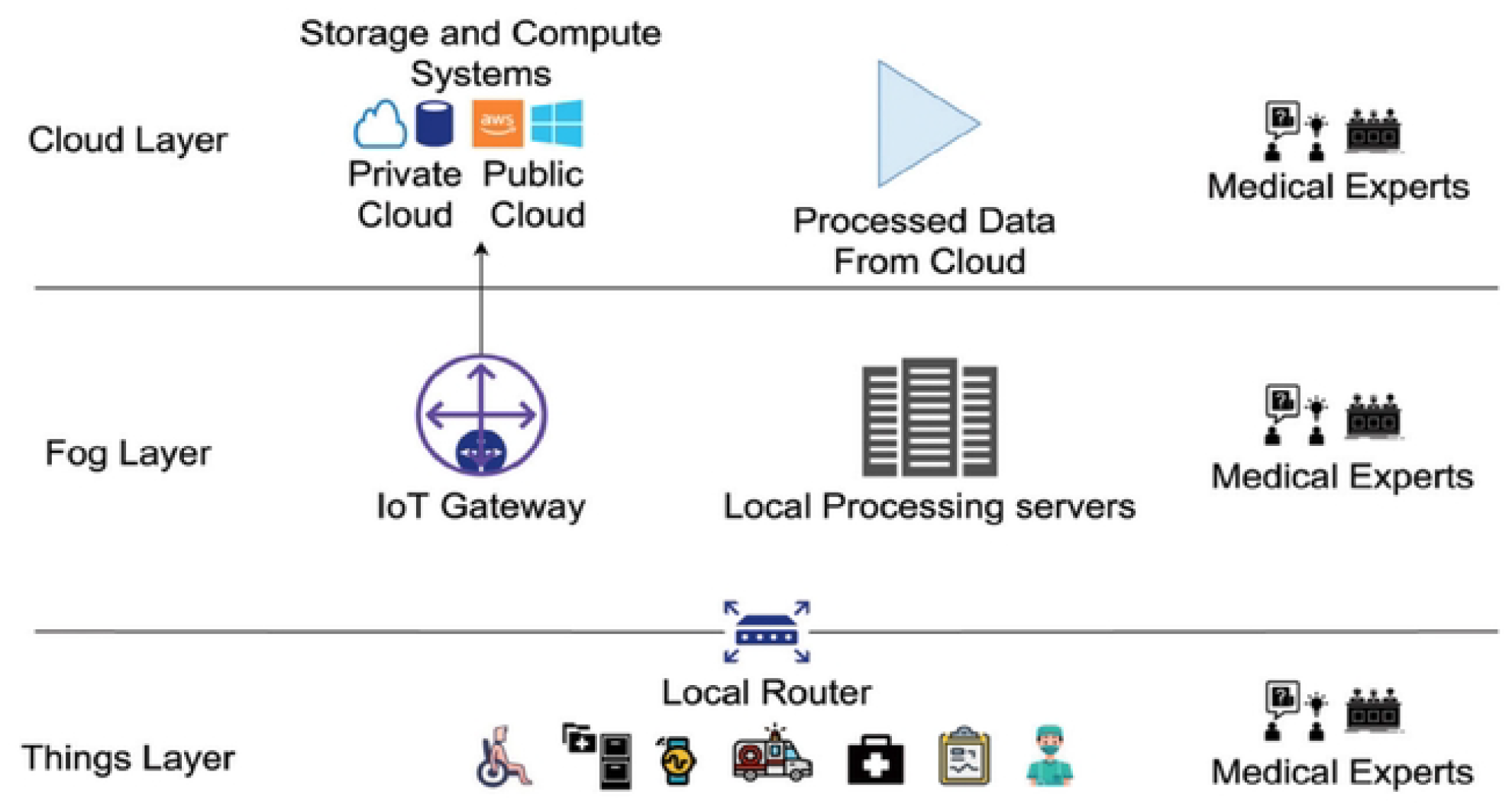
Overview of IoMT architecture [13].

- The ***thing layer*** in IoMT involves tracking of patient’s devices, sensors, and medical records communicating with users, obtaining data from wearables, monitoring systems, and remote care. Devices are securely placed and linked to the fog layer for processing. Data is analyzed at both the fog and cloud levels to yield significant insights. Healthcare practitioners use the router to access patient data in real time with minimum latency.
- For sparsely distributed fog networking, the ***fog layer*** links the cloud and objects layers with local servers and gateways. Lower-layer devices allow for real-time replies, whereas servers provide data security and integrity. Gateways send data to the cloud to be processed. This layer allows healthcare practitioners to access patient data in order to minimize delays and assure timely access.
- The ***cloud layer*** includes data storage as well as computational resources for data analysis and decision-making systems. It offers a comprehensive framework for integrating big medical and healthcare systems, therefore simplifying their day-to-day operations. This layer is in charge of storing data generated by healthcare systems and performing appropriate analytical operations for future applications.

## 4 Related Work

As IoMT systems are dynamic in nature, they frequently face challenges due to concept drift, which occurs when data distribution changes over time. The performance of model learning can be adversely affected by this phenomenon. To solve this issue, several solutions for drift detection and adaptation have been proposed. These strategies are designed to address specific concept drifts, perform effectively in a variety of circumstances, and are relevant to our proposed framework to varying degrees. This section explores the various techniques that have been proposed for building an IoMT system. Several examples of these techniques are presented, with an emphasis on analyzing existing approaches for detecting and adapting to concept drift within the IoMT framework.

In a prior study [15], a smart monitoring system leveraging the Internet of Medical Things (IoMT) was introduced to aid individuals exposed to COVID-19. Constructed using the MATLAB tool and employing the Mamdani fuzzy inference approach, this system gathers user data related to COVID-19, assesses their health status, and provides recommendations on whether consulting a professional for quarantine is advisable. The reported accuracy of this proposed system is approximately 83%. Similarly, another initiative [16] suggests an Internet of Things (IoT)-based system designed to gather real-time symptom data from users, aiming to detect potential COVID-19 cases early, monitor treatment responses, and analyze relevant data. Comprising Symptom Data Collection, Quarantine/Isolation Centre, Data Analysis Centre (employing machine learning techniques), Health Physicians, and Cloud Infrastructure, the study evaluated eight machine learning algorithms on an authentic COVID-19 symptom dataset, with five of them achieving accuracy rates exceeding 90%.

Recently, wearable gadgets have made important contributions to the advancement of health monitoring systems and the growth of IoMT, assisting in early disease identification and lowering death rates. The research [17] focuses on the application of machine learning techniques to forecast cardiac illness and presents an IoMT framework based on MSSO and ANFIS. The model outperforms previous techniques with an outstanding accuracy of 99.45% and a precision of 96.54%. Moreover, this study [18] used IoT and a support vector machine (SVM) to develop a technique for predicting cardiac illness. The suggested method predicted heart illness with 97.53% accuracy by collecting cardiovascular data such as blood pressure, body temperature, and heartbeat via IoT devices. Large data quantities, on the other hand, have an impact on accuracy.

### 4.1 Concept Drift Defination

An IoMT medical data stream consists of data instances that are continually changing and arriving sequentially [19]. In medical data streams, newly received data instances may have a pattern that differs from the preceding data. This occurrence is known as concept drift [20]. The Internet of Medical Things (IoMT) is a network of interconnected smart medical gadgets and applications that work together to improve smart healthcare. The continuous generation of streaming data from these devices raises concept drift challenges affecting model learning performance [21]. As stated in [21], Concept drift refers to a phenomenon observed in medical data streams, wherein the combined probability distribution *Prob*_*t*_(*X,y*) of the input variable *X* and its corresponding class data *y* ∈ *Y* undergoes notable changes between successive time intervals, *t* and *t* + 1 can be denoted by [22]:

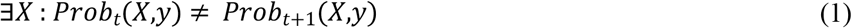

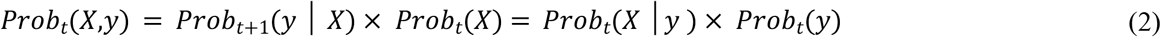

There are two categories of concept drift that exist: virtual drift and real drift. These distinctions are demonstrated in Figure 3(a). *Virtual drift* takes place when the marginal distribution *Prob*(*X*) alters but the subsequent distribution *Prob*(*y*│*X*) remains unchanged. This means that *Prob*_*t*_(*X*) is not equal to *Prob*_*t*+1_(*X*), but *Prob*_*t*_(*y*│*X*) is still equal to *Prob*_*t*+1_(*y*|*X*). Since changes in *Prob*(*X*) have no effect on categorization decisions, this kind of drift is known as virtual drift [23].

**Figure 3(a).**
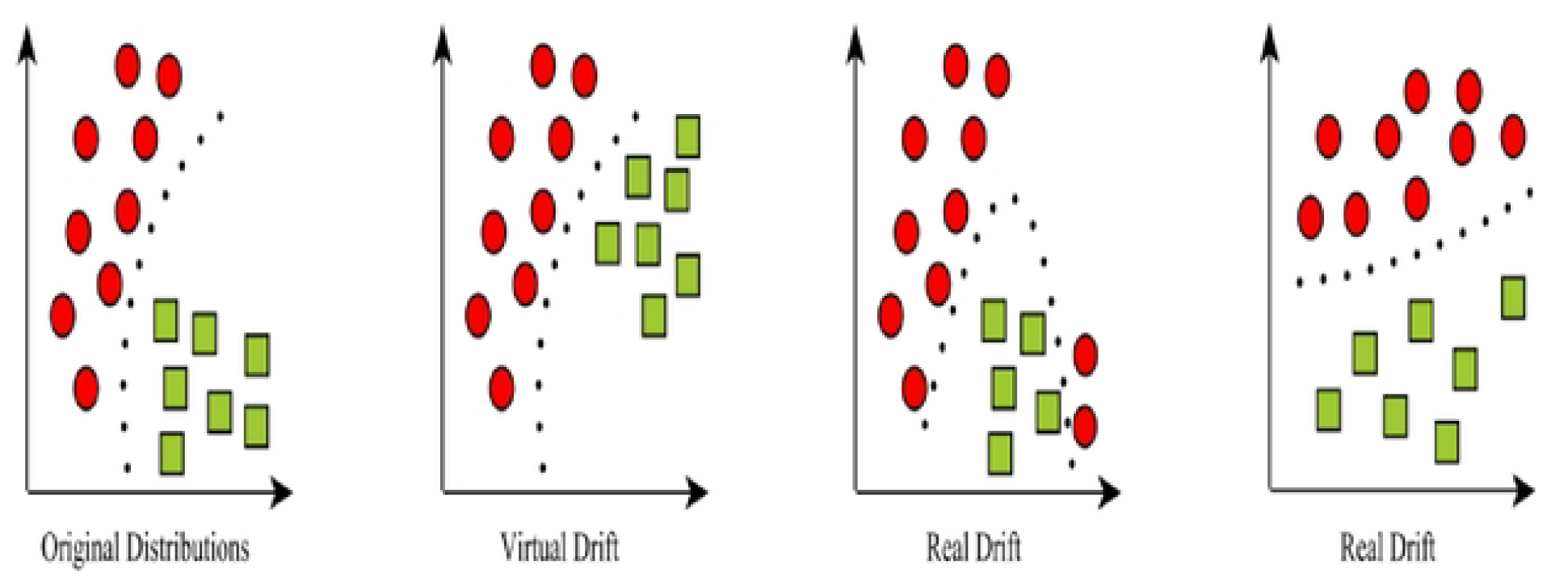
Visual representations of virtual and real drift derived from the initial distributions.

When concept drift influences the decision boundary, it’s termed as *real drift*. In the initial form of real drift, the alteration occurs in the subsequent distribution *Prob*(*y*│*X*), while the marginal distribution *Prob*(*X*) remains consistent. This indicates that *Prob*_*t*_ (*X*) remains equal to *Prob*_*t*+1_(*X*), while *Prob*_*t*_(*y*│*X*) diverges from *Prob*_*t*+1_(*y*|*X*). Such a form of drift, influencing the decision boundary, can result in a decline in the accuracy of the learning process.

Another type of real drift is a hybrid of the prior two, including changes in both the marginal distribution *Prob*(*X*) and the subsequent distribution *Prob*(*y*│*X*). In this case, *Prob*_*t*_(*X*) varies from *Prob*_*t*+1_(*X*), but *Prob*_*t*_(*y*│*X*) does not align with *Prob*_*t*+1_ (*y*|*X*).

Moreover, concept drift can be classified into four distinct categories, namely: (i) sudden/abrupt drift, (ii) gradual drift, (iii) incremental drift, and (iv) recurring drift. Figure 3(b) illustrates the various categories of concept drift.

**Figure 3(b).**
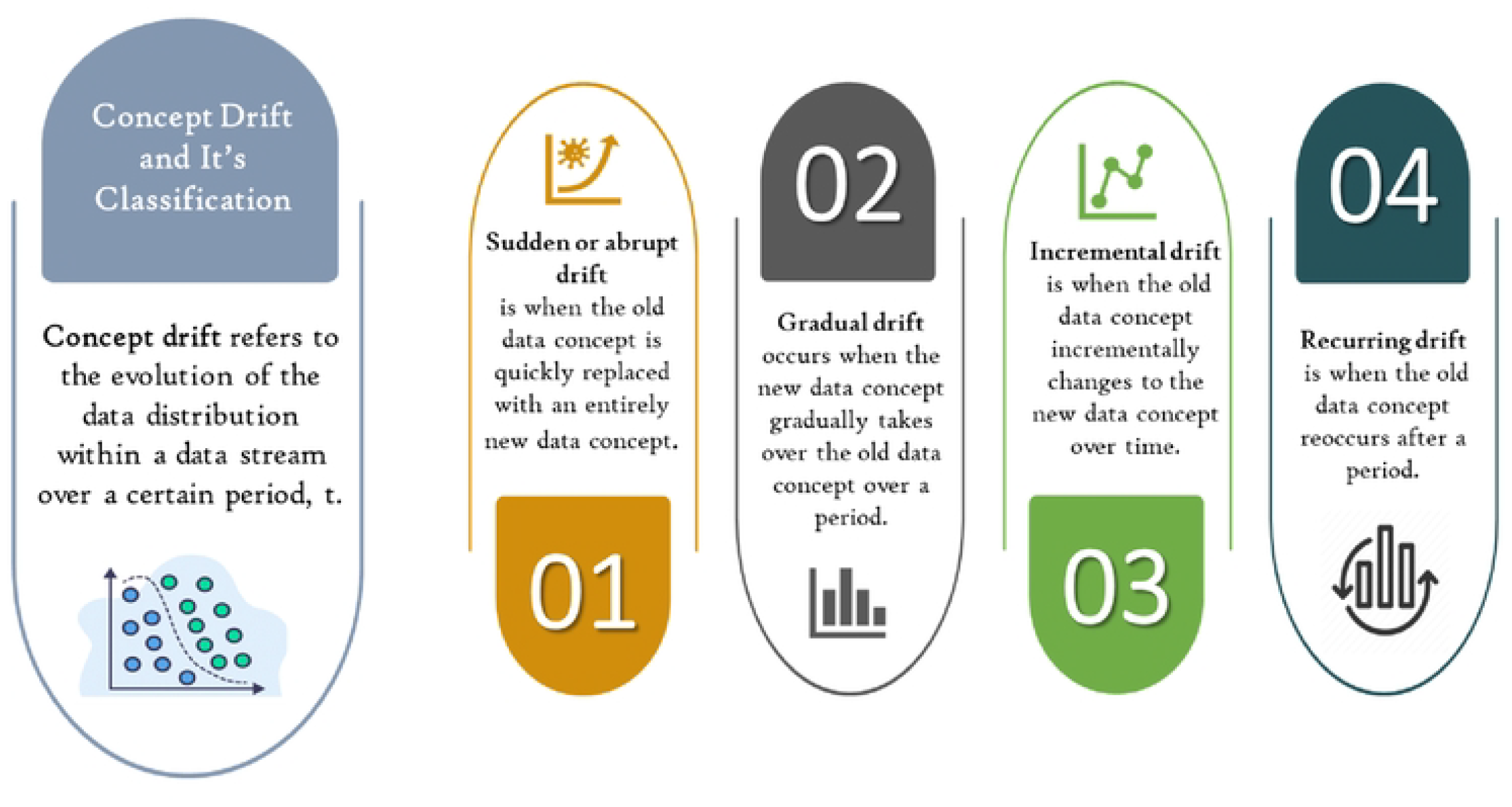
Types of concept drift [21].

*Sudden / Abrupt drift* happens when an event occurs inside a very small window size, causing an immediate fall in learned performance. *Gradual drift* can occur over a wide range of window sizes and is characterized by frequent shifts between two notions. *Incremental drift* is characterized by a linear shift that results in a gradual drop in learned performance. When an old data notion emerges after a certain period of time, this is referred to as *recurring drift*.

### 4.2 Concept Drift Detection Techniques

Concept drift detection is the term for the methods and procedures used to detect significant changes and measure concept drift. Evaluation of IoMT streaming frequently confronts concept drift issues due to the non-stationary IoMT environments, where the data distributions evolve over time. The existence of concept drift problems frequently decreases the performance and effectiveness of IoMT models and leads to serious security problems [24]. In general, techniques for identifying concept drift generally include four important steps. The initial stage involves data retrieval, encompassing diverse techniques for sampling data from the flowing data stream. Subsequently, the data modelling stage involves creating a representation of both past and current data distributions. Following that, the third step focuses on comparing and quantifying differences between historical and contemporary data distributions. Lastly, the fourth stage comprises performing a hypothesis test to assess whether the measured dissimilarity exceeds a preset level of significance as presented in Figure 4. Drift detection techniques are broadly categorized into two primary groups: distribution-oriented methods and performance-oriented methods.

**Figure 4.**
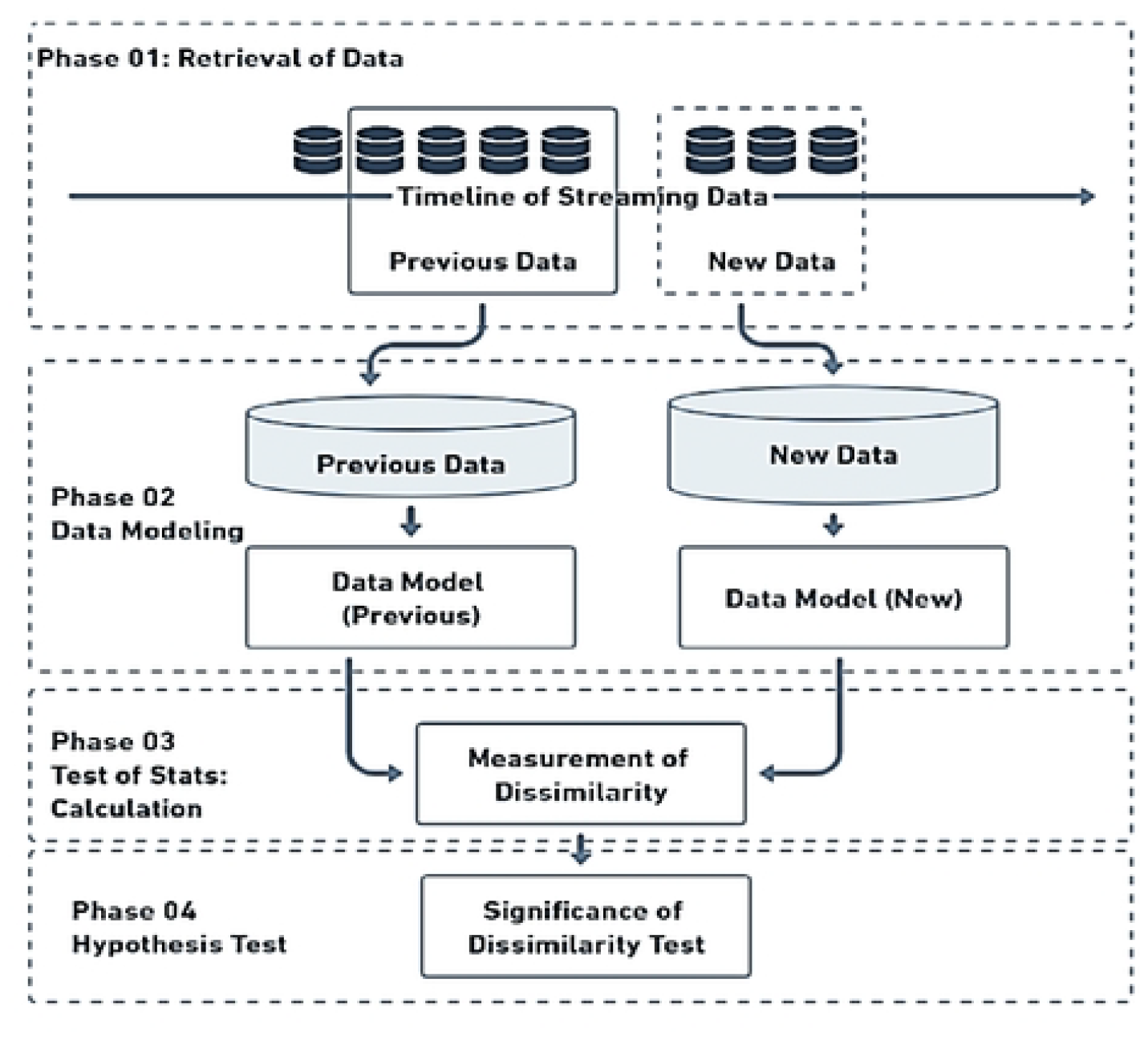
A general framework for concept drift detection.

#### 4.2.1 Distribution-oriented Concept Drift Detection

The design and implementation of distribution-oriented drift detection techniques involve evaluating and distinguishing data distributions from previous and more recent data sets within specified time frames. Substantial shifts in data distribution frequently serve as indicators of concept drift, prompting the need for model adjustments [25]. Various techniques can be employed to quantify data distributions, encompassing approaches like calculating the mean, measuring divergence, assessing information entropy, and utilizing metrics such as the Kullback-Leibler (KL) divergence.

ADWIN, introduced by [26], dynamically adjusts its window size (w) based on prediction outcomes to detect concept drifts. It evaluates two large sub-windows, expanding during stability and contracting upon drift detection. This allows for separate averages, revealing distribution differences. After drift detection, items gradually exit the window until no significant distinction remains. Drift detection in ADWIN2 occurs when the average distribution divergence between consecutive windows exceeds a threshold. It focuses on detecting gradual shifts over an extended period, reducing both time and memory complexity to O(log WS) for window size WS [27]. ADWIN demonstrates effectiveness in scenarios involving data streams characterized by gradual drift, owing to its ability to enlarge the sliding window to include a wide range for capturing prolonged shifts.

Moreover, OCDD, a one-class classifier-based technique for concept drift identification [28], employs a static one-class classifier to monitor outlier proportions in data windows as drift indicators. In the STEPD context, significant disparities between recent and older window examples prompt warnings and drift identification. STEPD parameters, with default values, include recent window size (w=30) and significance levels for drift detection (0.003) and warnings (0.05) [29].

The recently introduced equal-intensity k-means [30] is a space partitioning method using a greedy approach to determine initial centroids for k-means in space splitting. Concept drift is assessed by comparing newly partitioned distributions to previous ones using the chi-square probability test. Another method, a semi-supervised approach using Kullback-Leibler distance [31], initializes the model with labels from the first data window. Concept drift is detected when the classifier’s average confidence deviates from the reference mean by three standard deviations in the current data window. KL divergence computation involves the negative summation of event probabilities in distribution P, multiplied by the logarithm of the ratio of the probability in distribution Q to that in distribution P.

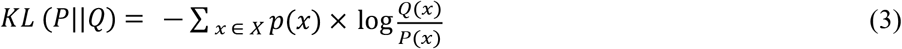

The “||” symbol signifies the concept of *“divergence”* or how distribution *P* diverges from distribution *Q*.

Distribution-oriented concept drift detectors address concept drift by monitoring shifts in data distributions. These methods visually represent evolving data distributions over time, offering enhanced accessibility. Distribution-oriented techniques excel at precisely identifying concept drift occurrences as they directly engage with the essence of concept drift. However, they reportedly entail higher computation costs compared to performance-oriented methods. In essence, distribution-focused techniques maintain a constant historical data window while concurrently shifting the recent data window to observe the phenomenon of concept drift.

#### 4.2.1 Performance-oriented Concept Drift Detection

Performance-oriented techniques monitor alterations in the prediction error rates of models to detect instances of concept drift [26]. The error rate of a model should demonstrate a progressive drop or remain relatively stable when more data samples are taken into consideration in situations where the data distribution is steady and free from drift. On the other hand, if the model’s error rate significantly raises as additional data is processed, this typically signifies the presence of concept drift.

The widely recognized performance-based concept drift detection algorithm, Drift Detection Method (DDM) [32], generates preemptive warning and subsequent drift signals. DDM starts by training a model with a set of samples, establishing an initial error rate as a benchmark. When a new instance arrives, the model’s prediction influences the cumulative error rate adjustment. If the cumulative error rate exceeds the warning threshold, DDM trains a new model using recent instances while using the existing model for predictions. Concept drift is signaled if the overall error rate significantly increases over the drift threshold, leading to the new learner replacing the old one for predictions. The landmark error rate is updated with the new learner’s error rate [33]. Although DDM performs well with abrupt drifts, its response time may be inadequate for timely detection of gradual drifts [34].

Several concept drift detection algorithms, such as Learning with Local Drift Detection (LLDD) [32], Early Drift Detection Method (EDDM) [35], Hoeffding Inequality-oriented Drift Detection Method (HDDM) [36], Fuzzy Windowing Drift Detection Method (FWDDM) [37], and Dynamic Extreme Learning Machine (DELM) [38], adopt approaches similar to Drift Detection Method (DDM). LLDD modifies error rate evaluation and hypothesis testing compared to DDM. EDDM, an enhanced version, uses similar mechanisms for drift and warning but struggles with gradual drift compared to distribution-based methods, and is susceptible to noise-induced false alarms. HDDM uses the Hoeffding inequality to locate significant drift zones. FWDDM improves data retrieval with a fuzzy time window, addressing DDM’s slow detection in gradual drift scenarios. DELM incorporates a foundational neural network while maintaining essential DDM techniques.

### 4.3 Concept Drift Adaptation Techniques

After drift detection, it is important to efficiently manage the detected drift so that the learning model is able to adapt to the newly discovered samples. As a solution to the drift adaptation challenge, three different methods can be used: adaptive algorithms, incremental learning, and ensemble learning.

#### 4.3.1 Adaptive and Incremental Learning Algorithms

Adaptive algorithms deal with concept drift by fully retraining or changing learning models in a modified data set after identifying drift. It is usually a combination of ML models and drift detection techniques. Non-stationary data stream analysis, particularly in IoMT, IoT, and Big Data, is receiving increasing interest. Existing algorithms face challenges with drift or reliance on task-specific information. To address this challenge [39] introduced the self-adapting memory with the k-nearest neighbor (SAM-KNN) approach, which employs a dual-memory method to store both new and old beneficial data to adapt current concepts and past concepts. KNN is used to train a learner in this algorithm.

Incremental learning is process to update the learning model incrementally, ? learn samples one at a time in chronological sequence. The Very Fast Decision Tree (VFDT) classifier is designed for high-speed data streams, learning incrementally without compromising space or time efficiency as instances increase [40]. It processes data streams in a single pass and achieves outcomes comparable to traditional machine learning techniques. However, VFDT doesn’t handle concept drift. To address this, the Concept-adapting Very Fast Decision Tree (CVFDT) variant is introduced. A Hoeffding tree (HT) is an incremental learning strategy that combines the Hoeffding constraint into a decision tree (DT) to efficiently adapt to input streams. This constraint allows the tree to adapt its structure progressively based on incoming input, allowing it to deal with dynamic and developing datasets [41]. The HT utilizes the Hoeffding constraint to determine the number of trials needed? for splitting, in contrast to a DT that selects the optimal split. Nevertheless, the HT does not possess mechanisms? to handle specific forms of drift. The Extremely Fast Decision Tree (EFDT) [42], formerly referred to as the Hoeffding Anytime Tree (HATT), varies from the HT in how it splits nodes. When it reaches the confidence threshold, the EFDT splits nodes immediately rather than waiting for the optimum split, which distinguishes it from the HT. Although the HT might perform better, the EFDT adjusts to? concept drifts more accurately because of its splitting method. Another incremental learning method, [43] Online Passive-Aggressive (OPA), adjusts to drift by? quietly responding to accurate predictions and firmly reacting to any failures. In contrast, SAM-KNN uses two memory modules to compensate for concept drift: Short-Term Memory ? (STM) holds the present concept, while Long-Term Memory (LTM) stores prior conceptions.

#### 4.3.2 Ensemble Learning and online Ensemble Learning Algorithms

Ensemble learning techniques have been proposed as a way to build effective learners for data stream analysis. These techniques can help to improve concept drift adaptation by combining the predictions of multiple learners. This can help to mitigate the effects of changes in the data distribution, as the ensemble will be less likely to be affected by any single change. Online ensembles and block-based ensembles are additional categories for ensemble techniques [44]. Three popular block-based ensembles are Accuracy Updated Ensemble (AUE), Accuracy Weighted Ensemble (AWE), and Streaming Ensemble Algorithm (SEA) [45].

Leverage bagging (LB) [46] is a fundamental ensemble online learning algorithm that builds and joins a number of base learners (such as HTs) using samples from the bootstrap and the majority voting method. Although LB is straightforward to build, it is vulnerable to data that is noisy. Utilizing this strategy, methods like SRP [47] and ARF [48] offer reliable and effective solutions to concept drift. ARF, for example, is an online ensemble algorithm that incorporates a number of HTs as base learners. By merging the predictions from several trees, it may gradually improve its classification performance. Unlike other methods, SRP uses a combination of online bagging and random subspaces to train its base learners. Online bagging entails training several base learners on different subsets of the data, whereas random subspaces imply selecting the subsets of features at random for each base learner. These approaches seek to diversify individual learners while enhancing the ensemble model’s overall performance. Due to the complexity involved in combining multiple models, these ensemble methods tend to have longer execution times despite exhibiting exceptional performance in adjusting to concept drift.

Although ensemble online learning approaches frequently outperform traditional learning approaches in the area of dynamic data stream analytics, they are frequently computationally costly. In order to enhance drift adaptation performance, this research proposes an online ensemble model that is stable and reliable.

## 5 Proposed Methodology for Concept Drift Adaptation and Detection

In this section, we provide an in-depth explanation of the proposed adaptive ensemble framework. Specifically, this framework introduces a novel ensemble approach that enhances the ensemble system’s diversity in a straightforward yet highly efficient manner.

### 5.1 Overview of the system

The architecture of proposed framework for detecting and adapting the concept drift in IoMT is shown in Figure 5. It has four main phases: dynamically preprocessing the IoMT data, adaptive feature selection using drift-based techniques, learning base models, and developing an online ensemble model. In the initial stage, data preprocessing techniques such as data balancing and normalization are employed to enhance the quality of the incoming IoMT data streams. Based on specific performance and efficiency requirements, ***an adaptive under-sampling or oversampling technique*** is automatically chosen. In addition, incoming IoMT data streams are automatically preprocessed by utilizing either dynamic min-max or z-score scaling approaches for data normalization.

**Figure 5.**
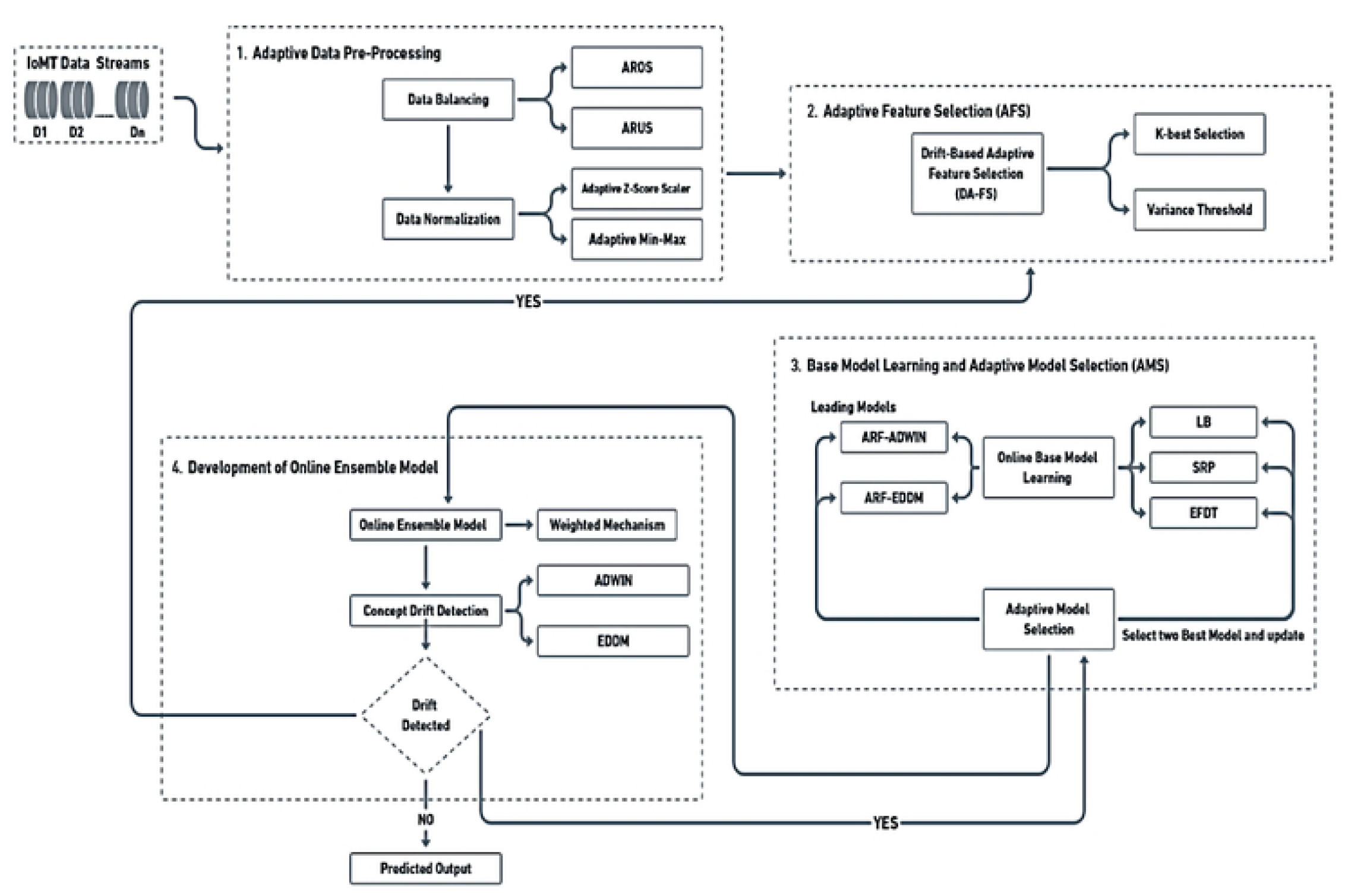
Architecture of Proposed Framework.

The second phase encompasses two pivotal steps: online base model learning and adaptive model selection (AMS). During the online base model learning stage, six base online learners are trained using preprocessed data streams. Specifically, for this study, we opted for two Adaptive Random Forest models, each incorporating a distinct drift detection mechanism: Adaptive Window (ADWIN) and Early Drift Detection Methods (EDDM). Designated as leader models, these two were selected. Additionally, we chose the top-performing models from four widely utilized online learning methods: extremely fast decision tree (EFDT), leverage bagging (LB), streaming random patches (SRP), and online passive aggressive (OPA). In the presence of concept drift, the adaptive model selection (AMS) module dynamically adjusts by reassessing and selecting appropriate base models that align with the updated data distributions. Subsequently, within a defined timeframe, these chosen learning models undergo updates with the incorporation of new concept instances. In the final stage, the proposed weighted method is employed to combine the outputs of the selected base learners for the online model ensemble, taking into consideration their prediction probabilities and real-time error rates.

### 5.2 Adaptive Data Preprocessing in the Context of Internet of Medical Things (IoMT)

Data preprocessing primarily seeks to improve the overall quality of data streams, with the ultimate goal of improving model learning performance. IoMT data streams present two possible issues related to data quality: imbalances in class distribution and variations in feature ranges. In order to resolve these issues, data sampling and normalization approaches can be employed. In dynamic IoMT systems, the distributions of classes and the ranges of features undergo continual changes, resulting in dynamic variables that exhibit significant fluctuations over time. Therefore, online data stream analysis should incorporate adaptable data preprocessing, as opposed to the static pipelines used in traditional machine learning.

#### 5.2.1 Data Balancing

As IoMT data streams undergo continuous evolution, achieving an equitable distribution of all classes in classification problems becomes challenging, giving rise to issues related to class imbalance. When addressing imbalanced data sets in training, there is the possibility of producing biased models with poor performance. To address challenges associated with class imbalance, techniques such as under-sampling and over-sampling can be utilized [49][50]. These strategies attempt to minimize the effects of unbalanced class distributions and improve the overall performance of the models. In order to attain balanced data, the adaptable random under-sampling (ARUS) method randomly removes data samples from the majority classes. Conversely, adaptable random over-sampling (AROS) maintains data balance by continuously generating additional samples for the minority classes [50].

The adaptable characteristics of ARUS and AROS allow for immediate adaptations to changes in class proportions within dynamic data streams. This ensures continuous maintenance of balanced data. ARUS, with its faster speed, is particularly well-suited for efficiency-focused IoMT systems, while AROS, prioritizing accuracy, proves more suitable for performance-oriented IoMT systems.

#### 5.2.2 Data Normalization

Machine learning heavily relies on data normalization. The advantages of this include faster algorithm convergence, improved model accuracy, and decreased impact from outliers. Z-score normalization normalizes data by subtracting the mean of the dataset from each value and dividing it by the standard deviation. By doing this, the dataset’s mean becomes 0 and its standard deviation becomes 1. The Z-score normalization technique is generally employed as a method of normalization. This method effectively regulates characteristics, enabling models to discover pertinent patterns more precisely. It is particularly effective at handling outliers in IoT data streams [51] [52].

The following formula is used to normalize the values in a dataset:

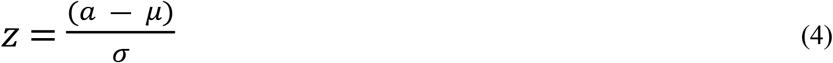

The variables in this equation are *z*, which stands for the Z-score, *a*, which stands for the starting point, *μ*, which stands for the dataset’s mean, and *σ*, which stands for its standard deviation.

Min-max normalization, also known as feature scaling, is a method used to preprocess data. It involves a simple linear transformation on the original data to ensure that all values are scaled within the range of 0 to 1. This technique is commonly applied to ensure uniformity and comparability of different features in the dataset, making it easier for machine learning algorithms to process the data effectively. The formula used to achieve this normalization is as follows:

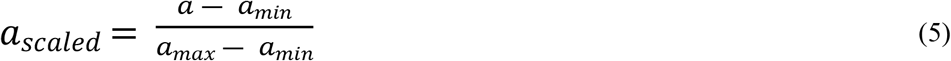

Where:

- *a* is the original data point.
- The variable *a*_*min*_ represents the smallest value observed for the feature within the dataset.
- The variable *a*_*max*_ denotes the highest value observed for the feature within the dataset.

Similarly to Z-score normalization, the ongoing minimum, and maximum values for each processed data sample in real-time are referred to as min and max. These values undergo continuous adjustments as additional data samples are processed, ensuring that normalized values for the processed samples can dynamically adapt to changes in data distribution. The primary objective of implementing this method is to effectively tackle issues related to concept drift detection within the realm of IoMT. Consequently, min-max normalization has been selected as the preferred normalization technique within the proposed framework.

### 5.3 Drift oriented Adaptive Feature Selection (DA-FS) method

The main objective of feature selection is to provide revised information with the most effective input characteristics to improve learning performance since the initial attributes are frequently not the most suitable ones for training an effective machine learning model. Additionally, the process of feature selection can also enhance the effectiveness of model learning by removing characteristics that are irrelevant or noisy. The approach introduced in this study, known as Drift-oriented Adaptive Feature Selection (DA-FS), employs a combination of two feature selection methods: variance threshold and select-k-best. The Variance Threshold acts as a feature selector, removing low-variance characteristics from the dataset that have no significant impact on the modelling process [53]. The sample’s variance is determined employing the following formula given a data sample with n observations;

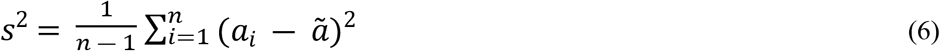

Here, *n* represents the count of analyzed samples, *a*_*i*_ stands for an input attribute, and 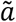 indicates the average value of *a*_*i*_. The variance threshold approach can be employed to help eliminate characteristics whose variance is less than a certain threshold. A small variance suggests that the associated attribute lacks informative value, as it maintains identical values across most data instances. The efficiency of model learning can be improved by excluding certain low-variance features. This approach can be applied to unsupervised learning models since it allows attribute selection without predetermined labels.

The select-k-best method is a well-known method for feature selection. In this process, the significance of each input attribute is determined by computing the correlation between that attribute and the target variable and selecting the top k attributes with the highest importance scores. The Pearson correlation coefficient can be used to derive feature importance scores, which are used to calculate the relevance of features. This frequently used measure, which quantifies the correlations between two variables, is expressed as follows:

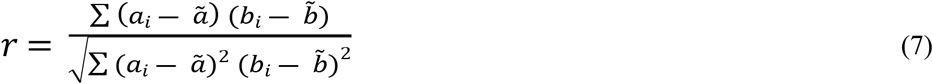

In this equation, *r* symbolizes the Pearson Correlation Coefficient. The variables, *a*_*i*_ represent input attributes, *b*_*i*_ stands for the target variable, and 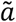 as well as 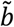 denote the mean values of the input attribute and target.

The value of ‘r’ can vary from ‘-1’ to ‘+1’. A value of ‘0’ denotes the absence of connection between the two variables. Indicating that a rise in one variable causes an increase in the other, a number greater than 0 denotes a positive correlation between the variables. In contrast, a value below ‘0’ denotes a negative correlation, implying that a rise in one measure corresponds to a drop in the other. Hence, this metric may quantify the significance of the association among each characteristic and the target variable, facilitating the evaluation and comparison of the significance of various attributes.

#### Algorithm 1

Drift oriented Adaptive Feature Selection (DA-FS)

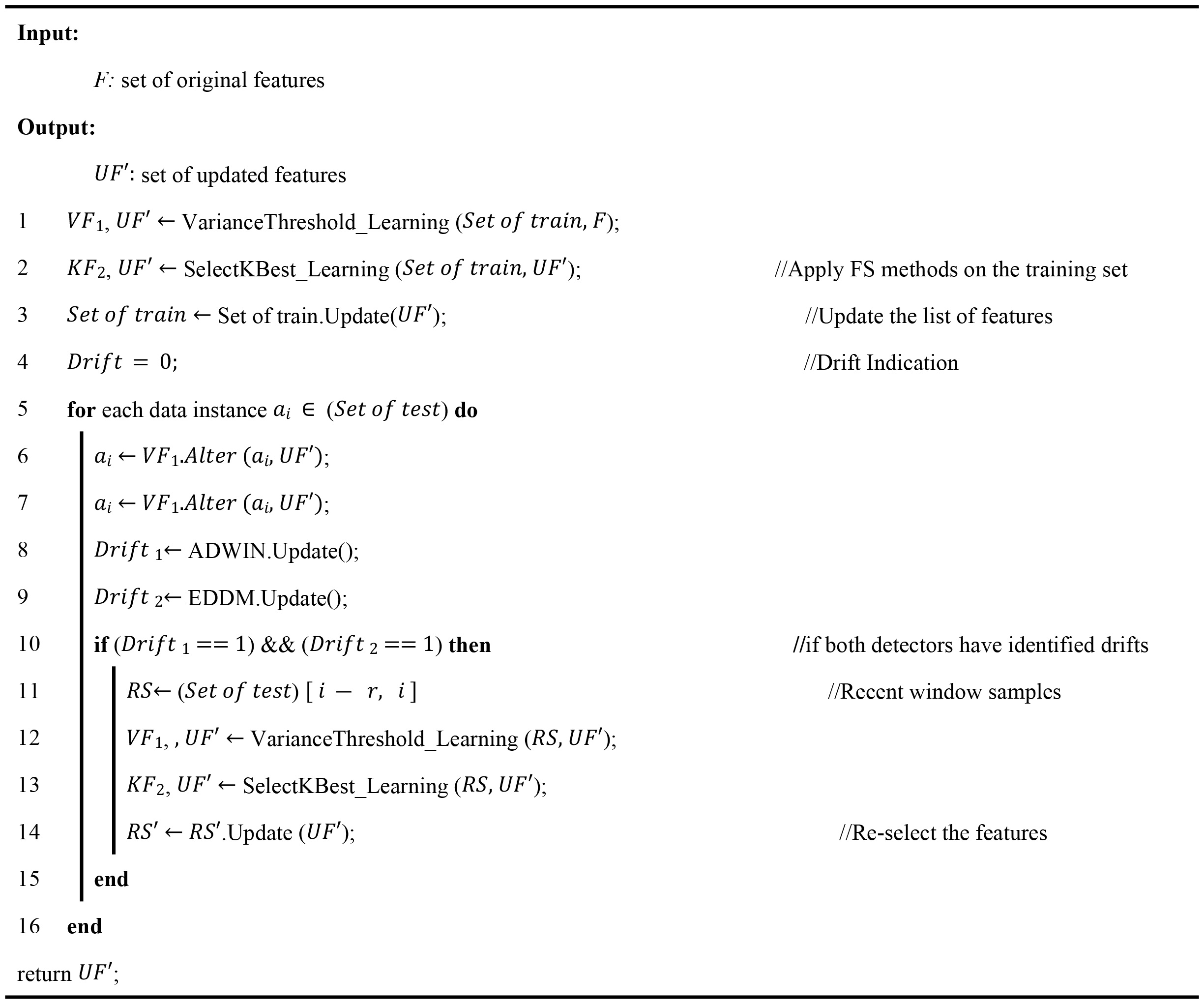

Algorithm 1 provides a detailed overview of the key steps in the DA-FS process. In the initial phase, the algorithm uses the variance threshold *VF*_1_, and the select-k-best method *KF*_2_, to extract the primary best possible attribute set *UF’*from the training dataset (*Set of train*). Subsequently, this initial *UF’*is employed to adaptively update the attribute list for each arriving data sample ai originating from the online test dataset (*Set of test*). Incorporating both ADWIN and EDDM drift detectors, an adaptive feature re-selection process is employed. When a drift is identified by ADWIN and EDDM, the variance threshold and select-k-best feature selection approaches are triggered. Subsequently, they undergo a process of relearning recent data samples within a defined time interval RS’, considering these as the novel concept samples. This relearning process results in the generation of an updated optimal feature set, denoted as *UF’*. To reduce the frequency of feature re-selection, the feature set update occurs only when both drift detectors signal the presence of concept drift. During the ongoing process of analyzing data streams in real-time, this method of selecting features in response to detecting concept drift is systematically executed whenever a concept drift event is detected. The underlying principle of DA-FS’s drift adaptation functionality is grounded in the idea that as concept drift occurs and data distribution shifts, the most appropriate feature set will also undergo changes.

### 5.4 Base Model learning and Adaptive Model Selection (AMS)

We employ adaptive and lightweight online learning methods for the purpose of learning the base model based on data streams. The initial step involves creating fundamental base learners using a limited-scale training dataset. During the construction of the base learners, two main drift detection methods (EDDM & ADWIN) are used along with state-of-the-art drift adaptation method ARF to construct a robust ensemble model [45]. In contrast to EDDM, which analyzes the standard deviation of the distance between classification errors and alerts to a potential drift when this distance rises, where ADWIN compares the standard deviation of data between two dynamically adjusted sliding frames. There are two base learners, namely ARF-ADWIN and ARF-EDDM the following are the reasons for selecting each of them:

All of the models mentioned are online ensemble models that are well-suited for adapting to concept drift in IoMT data streams. They are all developed with several HTs, which are effective incremental learning base models.

- The Adaptive Random Forest (ARF) [48] stands out as a proficient approach to online learning. This approach, which is based on Hoeffding Trees (HT) [41], offers a substantial benefit since it can effectively and reliably analyse data streams in a variety of circumstances.
- The Adaptive Random Forest demonstrates that it can effectively manage changes in data streams while maintaining reliable prediction ability. This is accomplished by combining an adaptive mechanism with several Hoeffding Trees to maximize their potential. Our framework employs ARF-ADWIN and ARF-EDDM as the fundamental classifiers to effortlessly incorporate the powerful features of ARF.
- While ADWIN is better at identifying gradual drifts, EDDM is more effective at spotting sudden drift. Both techniques are combined in the suggested ensemble model to efficiently find both kinds of drift.
- As one of the major benefits of the selected models, ARF-ADWIN and ARF-EDDM is their capability to provide robust performance with shorter execution times.
- In order to increase the potential for learning performance improvements, an effective ensemble model has to ensure a wide range of unique foundational learners. On the other hand, an ensemble with little variation frequently exhibits performance closely resembles that of a particular foundational learner.

### 5.5 Online Ensemble Framework for Drift Adaptation: AEF-CDA

The study introduces an Adaptive Ensemble Framework for Concept Drift Adaptation (AEF-CDA) for integrating foundational learners into IoT data stream analytics. In contrast to the fixed or predefined weights frequently employed in numerous existing ensemble techniques, AEF-CDA dynamically assigns weights to the foundational learners, modifying them in real-time in accordance with their performance. The target class predicted using can be expressed by the following mathematical equation when a data stream *D* = { (*a*_1_, *b*_1_),……(*a*_*n*_, *b*_*n*_) }, is taken into consideration, and the target variable has *c* distinct classes, *b* ∈ 1,. ….,*c*, for each input *a*.

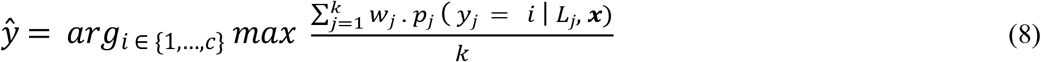

According to the suggested framework, *L*_*j*_ stands for a foundational learner, where *k* denotes the total number of foundational learners, and in this case, *k* equals 4. Additionally, *p*_*j*_ (*y*_*j*_ = *i* | *L*_*j*_, ***x***) signifies the prediction probability of class value *i* for data sample *x* when utilizing *L*_*j*_ as the jth foundational learner. Lastly, *w*_*j*_ represents the weight assigned to each base learner *L*_*j*_. The real-time rate of error is determined after the data samples have been analyzed by splitting the total number of analyzed samples by the number of samples that were erroneously classified. The weight assigned to each foundational learner, abbreviated as *w*_*j*_ is determined by finding the reciprocal of its associated real-time rate of error, which is denoted as:

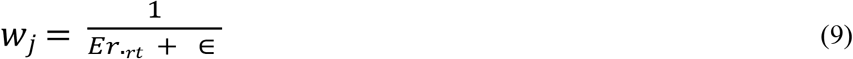

where ∈ serves as a minor constant to prevent the number in the denominator from being zero. If base learner’s *Er*._*rt*_ →0 for a, it is given a very high weight of 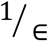, as it is already capable of making excellent predictions.

The weighting function in consideration can be thought of as an enhanced variant of the weighting function employed in the AUE approach. To enhance the precision of drift adaptation, the AEF-CDA model, as proposed, opts for the utilization of real-time error rates for computing base learner weights. Typically, the complexity of base learners plays a pivotal role in determining the time complexity of ensemble models. Given the relatively small values of *n* (the number of *c* class values in the target variable) and *k* (the number of base learners), AEF-CDA boasts a time complexity of merely *O*(*nck*).

In order to achieve effective drift detection and adaptation, the proposed AEF-CDA framework incorporates two foundational online learners including: ARF-ADWIN and ARF-EDDM. These two models have been designated as the leader models. Furthermore, we chose the two best performing models from four extensively used online learning methods: extremely fast decision tree (EFDT), leverage bagging (LB), streaming random patches (SRP), and online passive aggressive (OPA). ARF-ADWIN and ARF-EDDM are also employed to build the foundational learners in the suggested framework since they are the most advanced, high-performing drift adaptation techniques. Moreover, to obtain an optimized ensemble model, Particle Swarm Optimization (PSO) is used for hyperparameter tunning [54]. In the context of HPO problems, particle swarm optimization (PSO) is the most widely used metaheuristic algorithm. PSO algorithms use interaction among particles to identify and upgrade the existing global optimal point in each iteration until the ultimate optimum is identified [55].

The suggested AEF-CDA approach offers the following benefits over previous ensemble methods:

- When there is a disparity in class, the AEF-CDA strategy performs exceptionally well. In such challenging-to-forecast states, the model might prioritize minority classes and perform better if it weights predictions according to their confidence probability.
- This proposed AEF-CDA uses a reciprocal weighting function, which surpasses existing ensemble methods including AUE.
- The proposed framework relies on the confidence probabilities provided by all base classifiers for each class, which is deemed more reliable and flexible adaptable compared to numerous other ensemble learning methods that employ strict majority voting. To prevent making less effective decisions, this framework takes into account the uncertainty associated with each base classifier’s prediction for each data sample.
- It is possible to maintain high predictive performance while reducing computational challenges using the ensemble model. Additionally, the ensemble model can be used to process large amounts of data quickly, making it ideal for applications like internet of medical things (IoMT) and real-time analytics.
- ARF can be changed with other, higher-performing drift adaptation techniques if they are developed in the future in order to build a more effective ensemble model utilizing the same AEF-CDA approach.

## 6 Performance Evaluation

### 6.1 Experimental Setup

The execution of this framework utilized the Python 3.8 programming language and was carried out in Google Colaboratory. The implementation involved extending the Scikit-Multiflow framework [56] on a machine equipped with an i5-8350U processor and 8 GB of memory, serving as the central server machine for IoMT. Table 3 provides the insights of dataset employed for experiments.

**Table 3.** Datasets used for experiments.

| Dataset Name | Source | Category | Availability | Dataset Description |
| --- | --- | --- | --- | --- |
| IoMT Dataset | WUSTL-EHMS-2020-IoMT | Streaming Data | Publicly Accessible | The WUSTL-EHMS-2020 dataset was created using a real-time Enhanced Healthcare Monitoring System (EHMS) testbed. This testbed collects both the network flow metrics and patients' biometrics due to the scarcity of a dataset that combines these biometrics. |
| Intrusion Detection Evaluation Dataset (CIC-IDS2017) | CIC-IDS | Streaming Data | Publicly Accessible | The Canadian Institute for Cybersecurity Intrusion Detection System 2017 (CICIDS2017) dataset has the most updated network threats. The CICIDS2017 dataset is close to real-world network data since it has a large amount of network traffic data, a variety of network features, various types of attacks, and highly imbalanced classes. |
| IoT Intrusion Dataset | IoTID | Streaming Data | Publicly Accessible | IoTID20 dataset was created by using normal and attack virtual machines as network platforms, simulating IoT services with the node-red tool, and extracting features with the Information Security Center of Excellence (ISCX) flow meter program |

The suggested approach is evaluated using three datasets. The first dataset, known as WUSTL EHMS 2020 Dataset for (IoMT) Cybersecurity Research [57]. The EHMS testbed is segmented into four components: medical sensors, gateway, network, and control with visualization. The flow of data initiates from the sensors affixed to the patient’s body, transmitting to the gateway. Subsequently, the gateway forwards the data to the server for visualization, facilitated by the switch and router. A potential threat involves the interception of this data before it reaches the server. To counteract such threats, the Intrusion Detection System (IDS) takes charge of capturing real-time network flow traffic, as well as detecting abnormalities in the patient’s biometric data. The dataset comprises of 44 features, encompassing 35 network flow metrics, eight biometric features related to patients, and one feature designated for labeling purposes.

IoTID20 dataset [58], was produced utilizing both legitimate and malicious IoT devices to gather data analysis on IoT network traffic. The analysis of flow-based and packet data yielded 83 network properties, which were then used to identify and detect cyberattacks. These elements include a wide range of information, including active and idle times, the length of flows, and the total number of packets delivered and received in both forward and backward directions.

CICIDS2017, provided by the Canadian Institute of Cybersecurity (CIC), is the name of the second dataset used in the study [59]. It contains the most recent cyberattack scenarios that were accessible at the time. The attack patterns contained within the dataset evolved over time as a result of the launch of numerous attacks throughout various time periods to create the CICIDS2017 dataset. In the CICIDS2017 dataset, this led to several instances of concept drift.

To assess the model, a representative subset of the IoTID20 dataset, comprising 6,252 records, and a representative sample from the CICIDS2017 dataset, comprising 28,303 records, were chosen through the utilization of the k-means clustering sampling technique. Both the datasets, IoTID20 and the CICIDS2017 datasets show a significant class imbalance. IoTID20 has a normal-to-abnormal instance ratio of 94% to 6%, but CICIDS2017 has an abnormal-to-normal instance ratio of 80% to 20%. This feature makes it possible to assess how well the model performs on imbalanced datasets.

### 6.2 Validation

In this study, we treated the datasets as binary datasets with two labels: “normal” and “abnormal” since the primary objective of concept drift detection and anomaly detection systems is to classify between normal states and cyber-attacks. Utilizing hold-out and prequential validations, the proposed framework is assessed. As a component of the holdout evaluation, the initial model training is carried out utilizing the initial 10% of the data, with the subsequent 90% of the data reserved for online testing [60].

Within the process of prequential validation, alternatively known as test-and-train validation, each input sample from the online test set serves dual purposes. Initially, it functions to test the learning model, and subsequently, it contributes to the training or updating of the model. Given the imbalanced nature of both datasets utilized in this study, the performance assessment of the proposed concept drift detection and anomaly detection framework incorporates five metrics: accuracy, precision, recall, f1-score, and execution time. These metrics serve as evaluative measures to estimate the framework’s effectiveness in identifying concept drift and anomalies within the datasets.

### 6.3 Results and Discussion

The efficacy and superiority of the AEF-CDA method in addressing concept drift within the realm of online learning are emphasized through the presented figures and tables. Specifically, Table 4 provide a comprehensive comparative analysis between the proposed AEF-CDA technique and other state-of-the-art online learning approaches. In the experimentation phase using the WUSTL EHMS-IoMT dataset, six instances of concept drift were identified, as illustrated in Figure 6.

**Table 4.** Comparison of performance evaluation on WUSTL EHMS-IoMT dataset.

| Methods | IoMT-WUSTL-EHMS Dataset |  |  |  |
| --- | --- | --- | --- | --- |
|  | Accuracy (%) | Precision (%) | Recall (%) | F-1 Score (%) |
| EFDT | 97.99% | 91.11% | 88.8% | 89.94% |
| SRP | 94.25% | 66.71% | 70.74% | 68.66% |
| OPA | 92.11% | 55.14% | 53.71% | 54.41% |
| ARF-ADWIN | 99.32% | 96.38% | 96.06% | 96.22% |
| ARF-EDMM | 99.57% | 97.89% | 95.17% | 96.50% |
| **Proposed IoMT Model** | **99.64%** | **99.39%** | **99.96%** | **98.57%** |

**Figure 6.**
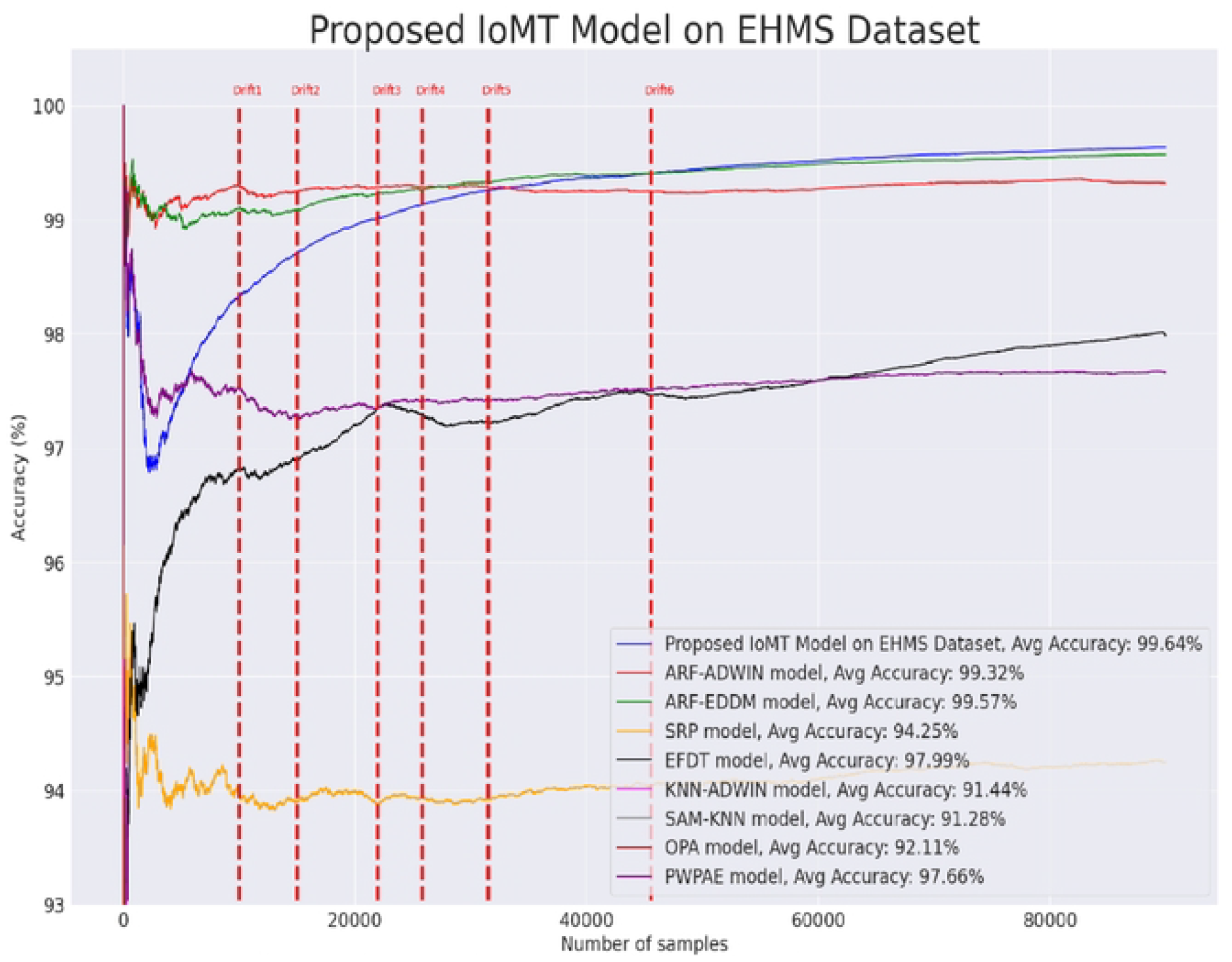
Proposed framework on WUSTL EHMS-IoMT dataset.

The results demonstrate that the AEF-CDA method, as introduced in this study, outperformed all other evaluated online learning models. It achieved the highest accuracy, reaching 99.64%, precision at 99.39%, recall at 99.96%, and an F1-score at 98.57% as shown in Figure 7. These findings underscore the superior performance of the AEF-CDA technique in effectively detecting and adapting to concept drift in online learning scenarios.

**Figure 7.**
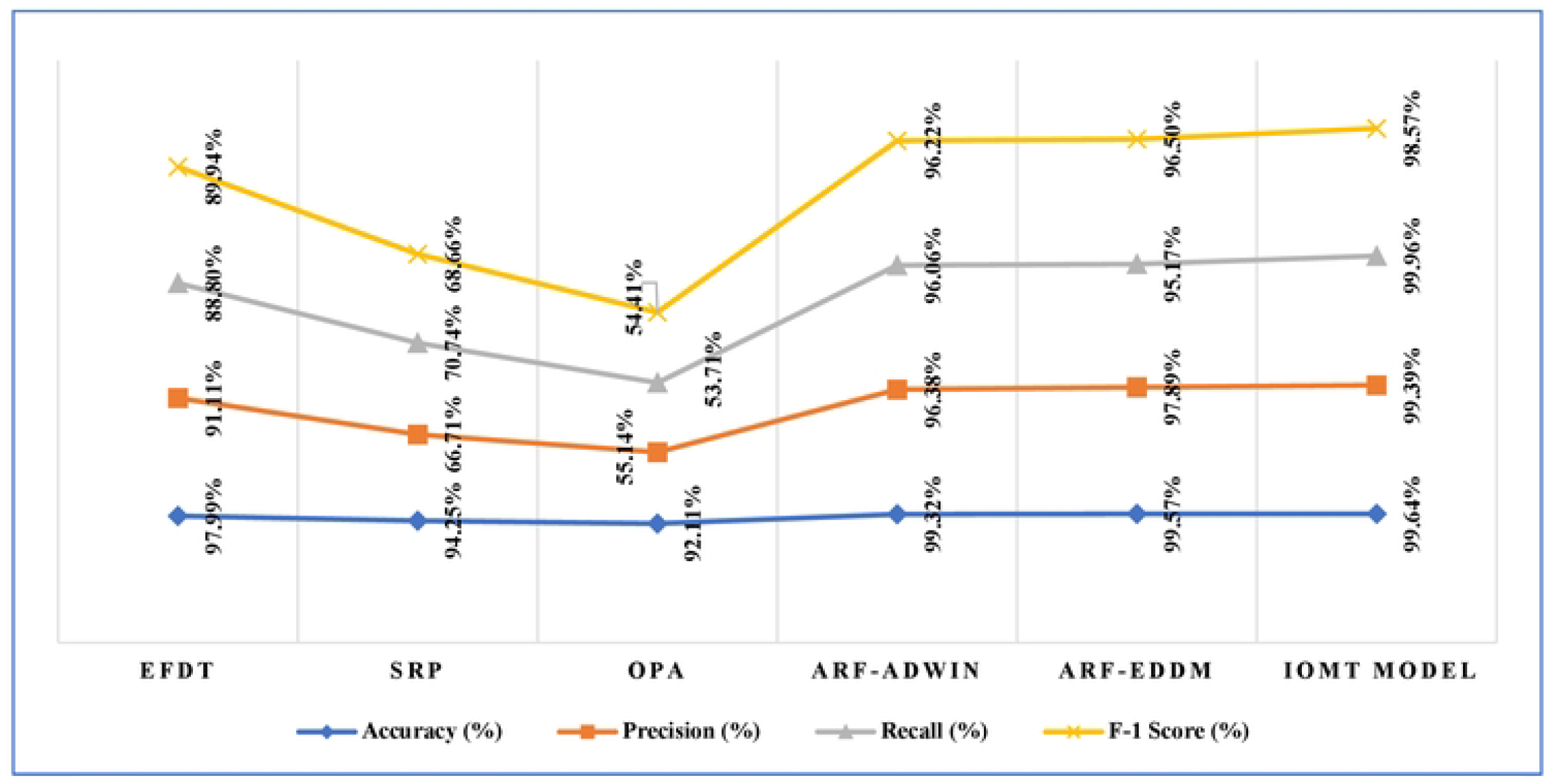
Comparative Analysis of proposed AEF-CDA model performance on WUSTL EHMS-IoMT dataset.

Based on the findings presented in Table 5, the proposed AEF-CDA approach showcases superior performance compared to all other models considered, with notable advantages in accuracy, precision, recall, and F1-Score. In the IoTID20 dataset experiment, two minor drifts were observed at the experiment’s onset, as depicted in Figure 8. All applied methods demonstrated the ability to promptly adapt and respond effectively, albeit with varying degrees of adaptability.

**Table 5.** Comparison of performance evaluation on IoTID20 dataset.

| Methods | IoTID20 Dataset |  |  |  |
| --- | --- | --- | --- | --- |
|  | Accuracy (%) | Precision (%) | Recall (%) | F-1 Score (%) |
| EFDT | 97.28% | 97.53% | 99.55% | 98.53% |
| SRP | 98.34% | 98.50% | 99.60% | 99.05% |
| LB | 98.32% | 98.09% | 99.50% | 98.77% |
| OPA | 91.84% | 95.99% | 95.27% | 95.63% |
| ARF-ADWIN | 98.06% | 98.50% | 99.68% | 99.09% |
| ARF-EDMM | 98.26% | 98.09% | 99.45% | 98.76% |
| **Proposed Model** | **99.14%** | **98.98%** | **99.81%** | **99.37%** |

**Figure 8.**
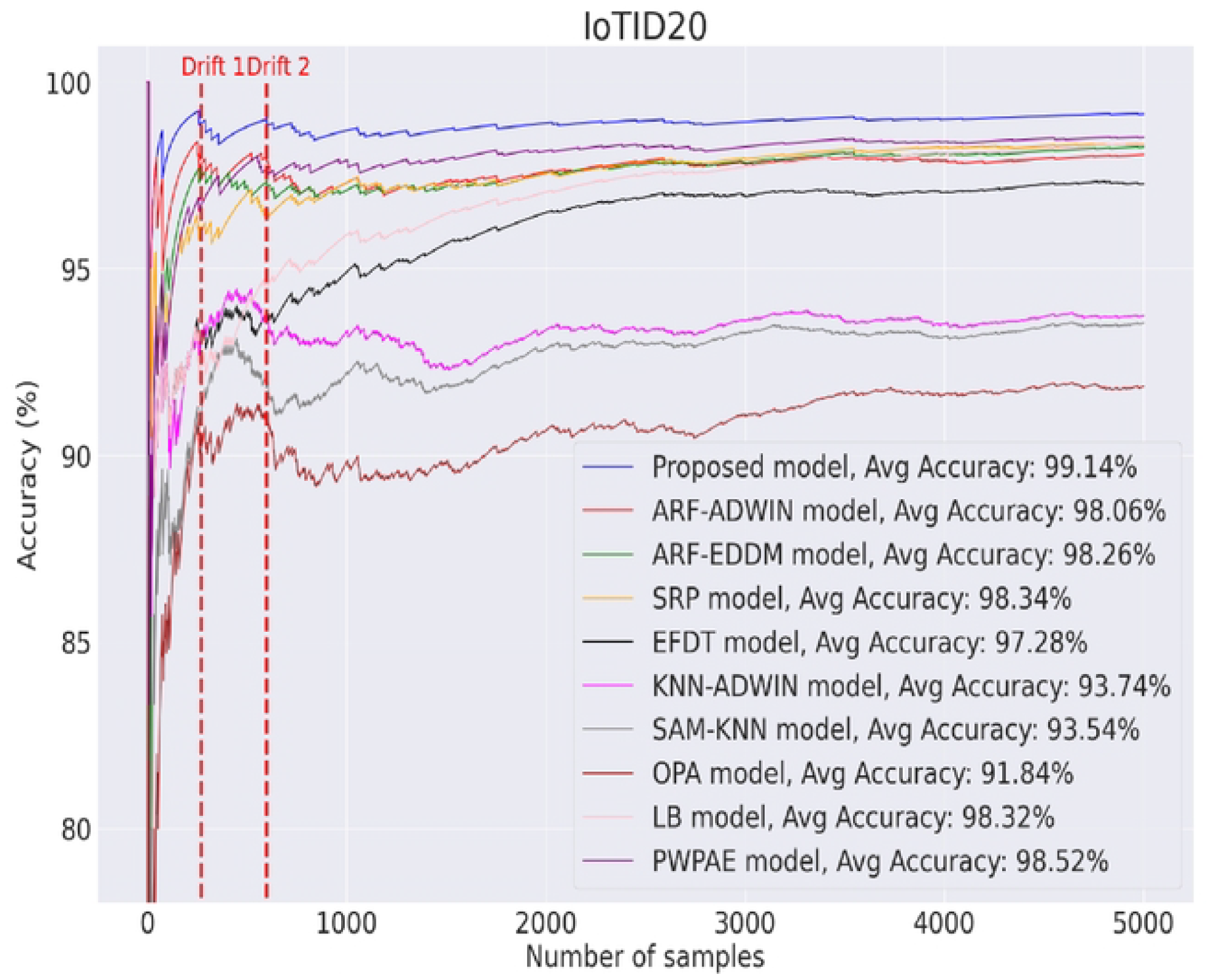
Proposed framework on IoTID20 dataset.

Notably, the AEF-CDA framework stands out among the assessed methods, achieving the highest accuracy at 99.14%, as illustrated in Figure 9. In contrast, when comparing AEF-CDA to the two fundamental learners (ARF-ADWIN and ARF-EDDM), their accuracy ranged between 98.26% to 98.06%. These results emphasize the superior adaptability and performance of the AEF-CDA approach in handling concept drift in the context of the IoTID20 dataset experiment.

**Figure 9.**
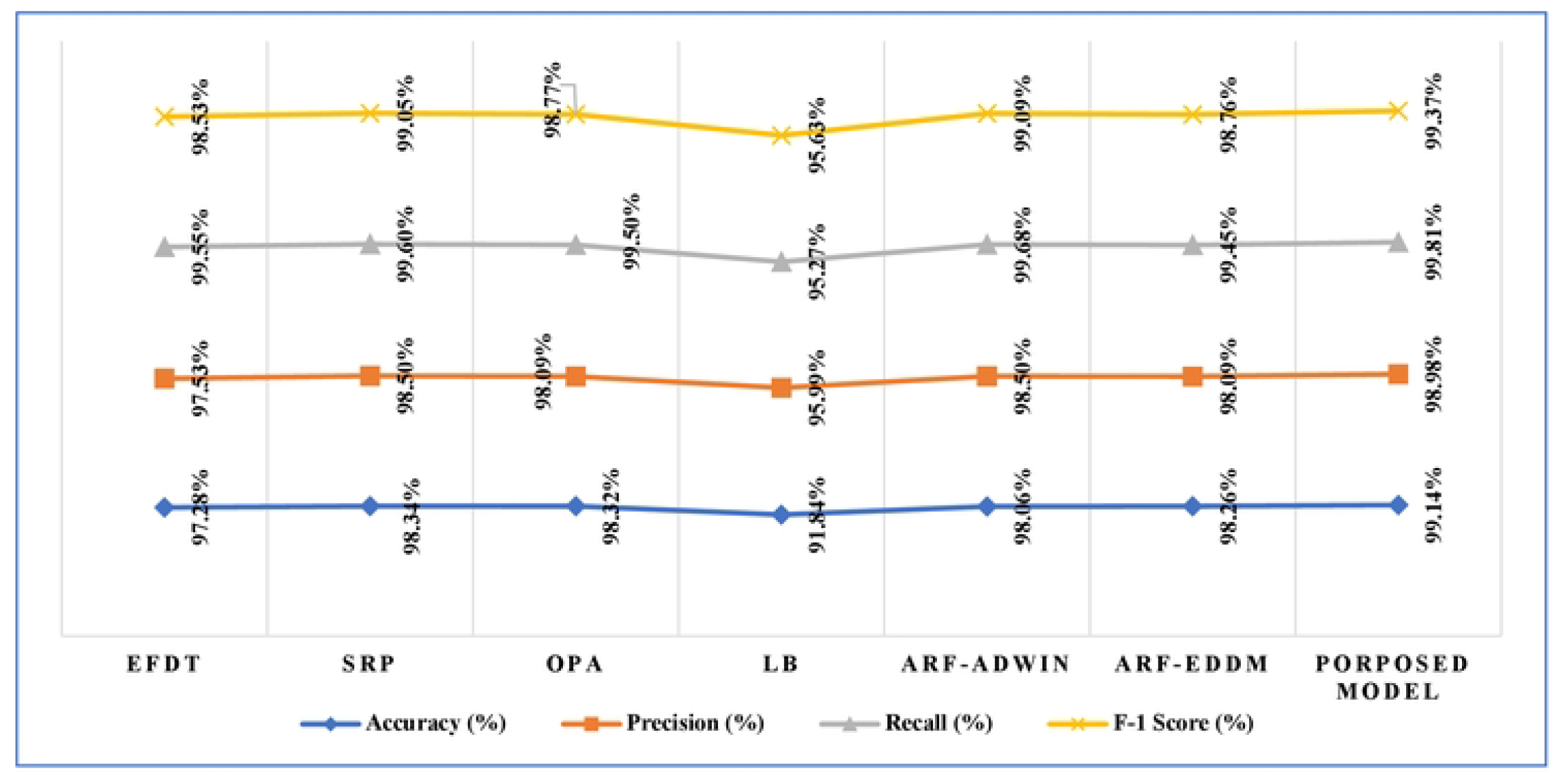
Comparative Analysis of proposed AEF-CDA model performance on IoTID20 dataset.

In the experimentation phase on the CICIDS2017 dataset, six instances of concept drift were identified, as depicted in Figure 10. Notably, the first three drifts (drifts 1, 3, and 5) were sudden, while drifts 2, 4, and 6 exhibited a gradual transition. Drawing insights from both the IoTID20 dataset and the proposed AEF-CDA method, foundational learners ARF-ADWIN and ARF-EDDM demonstrated swift adaptability to drifts, maintaining high accuracy in the CICIDS2017 dataset.

**Figure 10.**
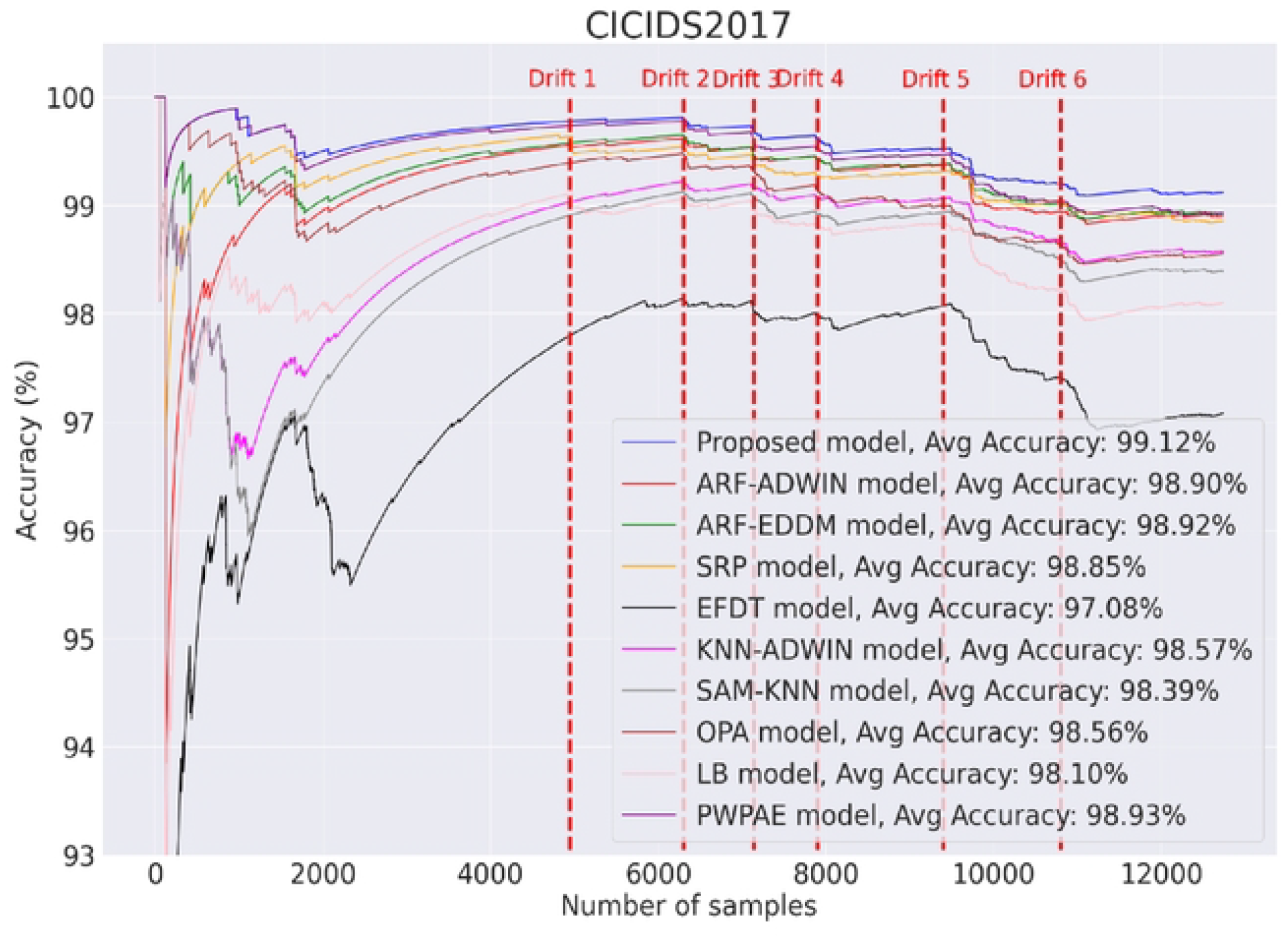
Proposed framework on CICIDS2017 dataset.

AEF-CDA emerges as the standout performer with a 99.12% accuracy rate, as illustrated in Figure 11 and supported by Table 6.This places AEF-CDA at the leading approach, confirming its position as the superior approach. Among the four follower models (OPA, SRP, EFDT, and LB), SRP and LB demonstrate notable proficiency. However, the accuracies of OPA, SRP, EFDT, and LB showcase a considerable range, spanning from 97.08% to 98.85%. These outcomes emphasize the effectiveness of AEF-CDA and highlight SRP and LB as particularly adept follower models in managing concept drift in the CICIDS2017 dataset experiment.

**Table 6.**
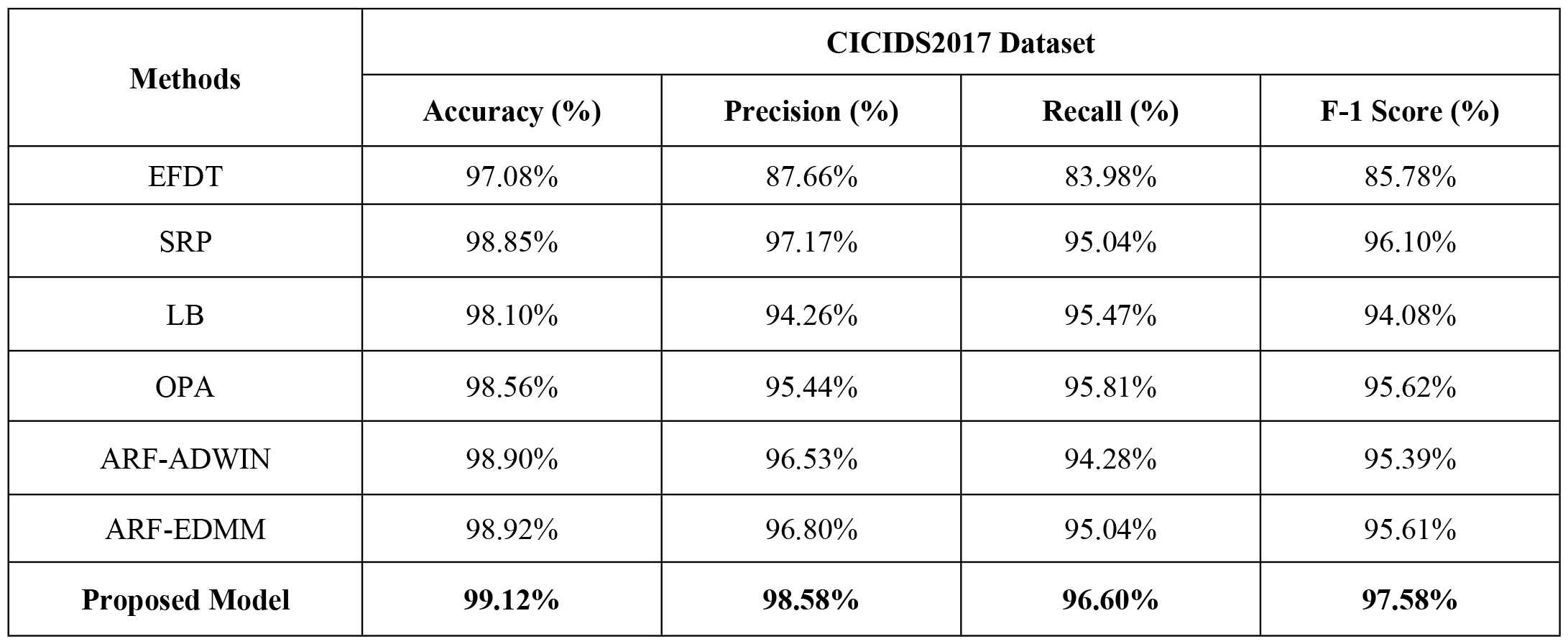
Comparison of performance evaluation on CICIDS2017 dataset.

**Figure 11.**
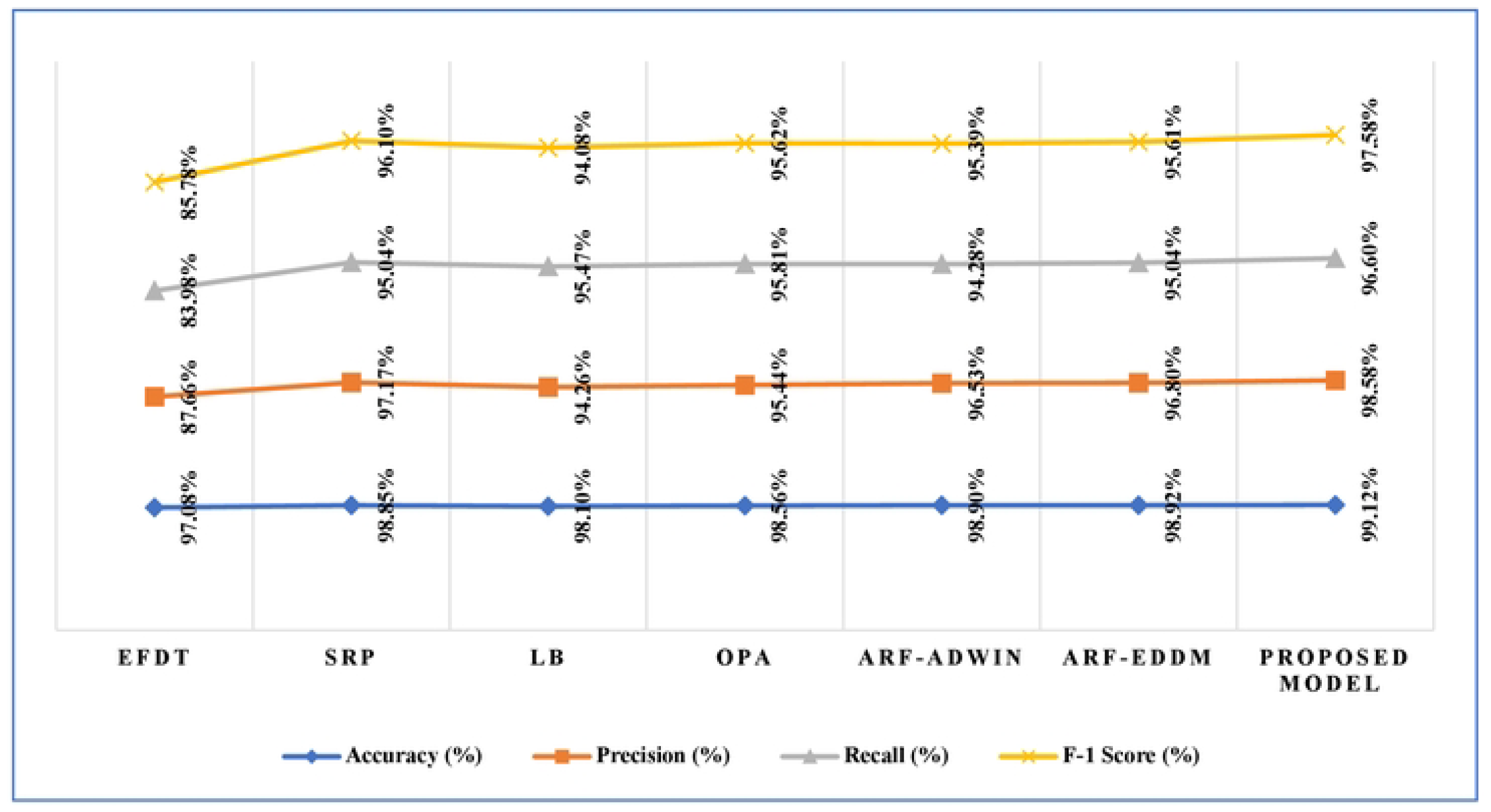
Comparative Analysis of proposed AEF-CDA model performance on CICIDS2017 dataset.

The suggested AEF-CDA approach uses min-max sampling to maintain high accuracy on a sampled subset despite the rapid generation of IoT data streams. With this approach, real-time data analytics are made easier, and the model learning speed is increased. Additionally, the SRP drift adaptation model can be replaced with another drift adaptation model with less complexity to reduce the average execution time of the proposed AEF-CDA approach. But when compared to other cutting-edge drift adaptation methods for IoT anomaly detection, the AEF-CDA approach now in use can obtain the best accuracy and F1-score.

## 7 Conclusions

The advancement of fifth generation (5G) wireless networks has significantly increased the use of Internet of Things (IoT)-based smart technologies, bringing people an enormous amount of comfort. However, as use has grown, there is also the risk of being a target of malicious cyberattacks increased. IoMT anomaly detection technologies have been developed to address this issue and protect IoMT systems from cyberattacks by examining IoMT data streams. There are many common characteristics associated with Internet of Medical Things (IoMT) data, most prominently their dynamic nature, occurring in non-stationary and rapidly changing environments, resulting in concept drift problems. This study presents a cutting-edge IoMT concept drift detection approach called AEF-CDA that successfully combats concept drift. The system uses a number of sophisticated drift adaptation algorithms to enable flexible modifications in a variety of situations. Our suggested approach demonstrates exceptional efficiency in dynamic IoMT and IoT data analytics, according to evaluation findings from one IoMT and two IoT datasets. It outperforms the performance of other great online learning algorithms, achieving accuracy of 99.64% on WUSTL EHMS-IoMT dataset, 99.38% on the CICIDS2017 dataset and 99.18% on the IoTID20 dataset. Moreover, the proposed framework can be extended to incorporate other drift adaptation methods with improved performance, diversity, and speed in future work.

Future directions for research could entail exploring the detection of drift types other than the common abrupt and gradual ones, such as incremental and recurring variations. However, a significant hurdle in pursuing these avenues is the scarcity of data streams showcasing these less typical drift patterns. Developing algorithms capable of effectively handling all forms of drift can be a complex undertaking. Additionally, another potential area of focus could involve tackling the challenges posed by partially or entirely unlabeled data streams in the context of addressing both concept drift and class imbalance issues.

## Data Availability

All relevant data are within the manuscript and its Supporting Information files.

https://www.cse.wustl.edu/~jain/ehms/index.html

https://www.unb.ca/cic/datasets/ids-2017.html

https://sites.google.com/view/iot-network-intrusion-dataset/home

## Notes

### Competing Interest Statement

The authors have declared no competing interest.

### Funding Statement

Yes

